# Imputation of structural variants using a multi-ancestry long-read sequencing panel enables identification of disease associations

**DOI:** 10.1101/2023.12.20.23300308

**Authors:** Boris Noyvert, A Mesut Erzurumluoglu, Dmitriy Drichel, Steffen Omland, Till F M Andlauer, Stefanie Mueller, Lau Sennels, Christian Becker, Aleksandr Kantorovich, Boris A Bartholdy, Ingrid Brænne, Julio Cesar Bolivar-Lopez, Costas Mistrellides, Gillian M Belbin, Jeremiah H Li, Joseph K Pickrell, Jatin Arora, Yao Hu, Boehringer Ingelheim – Global Computational Biology and Digital Sciences, Clive R Wood, Jan M Kriegl, Nikhil Podduturi, Jan N Jensen, Jan Stutzki, Zhihao Ding

**Affiliations:** Global Computational Biology and Digital Sciences (gCBDS), Boehringer Ingelheim Pharma GmbH & Co. KG, Biberach an der Riss, Germany; BI X GmbH, Ingelheim am Rhein, Germany; Department of Neurology, School of Medicine, Technical University of Munich, Munich, Germany; Gencove, New York, USA; Discovery Research, Boehringer Ingelheim Pharma GmbH & Co. KG, Ingelheim am Rhein, Germany; Institute of Cancer and Genomic Sciences, University of Birmingham, B15 2TT, UK

**Author notes:** These authors contributed equally to this work (Boris Noyvert, A Mesut Erzurumluoglu, Dmitriy Drichel, Steffen Omland, Till F M Andlauer). Joint senior authors (Jan Stutzki, Zhihao Ding). The full list of **Boehringer Ingelheim – Global Computational Biology and Digital Sciences** authors are listed in the ‘Author contributions’ section.

## Abstract

Advancements in long-read sequencing technology have accelerated the study of large structural variants (SVs). We created a curated, publicly available, multi-ancestry SV imputation panel by long-read sequencing 888 samples from the 1000 Genomes Project. This high-quality panel was used to impute SVs in approximately 500,000 UK Biobank participants. We demonstrated the feasibility of conducting genome-wide SV association studies at biobank scale using 32 disease-relevant phenotypes related to respiratory, cardiometabolic and liver diseases, in addition to 1,463 protein levels. This analysis identified thousands of genome-wide significant SV associations, including hundreds of conditionally independent signals, thereby enabling novel biological insights. Focusing on genetic association studies of lung function as an example, we demonstrate the added value of SVs for prioritising causal genes at gene-rich loci compared to traditional GWAS using only short variants. We envision that future post-GWAS gene-prioritisation workflows will incorporate SV analyses using this SV imputation panel and framework.

## Introduction

Human disease associations of single nucleotide variants (SNVs) and short insertions and deletions are routinely identified in genome-wide association studies (GWASs)^1,2^. By contrast, large structural variants (SVs) of >50 base pairs (bp) are typically neglected, despite functional roles in the context of disease. Each human carries 23,000-31,000 SVs^1^, often overlapping protein-coding genes or regulatory regions, thus enabling fine-mapping causal genetic variants^2,3^. Studies on single populations have already demonstrated the value of sequencing SVs for identifying causal variants underlying disease associations^4,5^.

Robust SV calling from traditional short-read sequencing is challenging because SVs are often longer than the average short-read length (**Figure 1a**)^3,6^. Long-read sequencing captures SVs reliably, but the high cost impairs its application to large-scale datasets. Reference panels constructed for imputation of SVs from genotyped samples enable biobank-scale genome-wide analyses of SVs. For example, imputing SVs in UK Biobank (UKB) can accelerate research on the genetic underpinnings of diverse diseases and facilitate the identification of novel therapeutic targets. To this end, we generated a publicly available multi-ancestry long-read sequencing-based SV imputation panel (**Figure 1b**). Simultaneously, an effort is ongoing to develop new methods for improved SV calling that utilises this dataset^7^. Another Oxford nanopore sequencing-based dataset of 1000 Genomes Project samples is being generated, SV calling of the first 100 samples was reported recently^8^.

**Figure 1.**
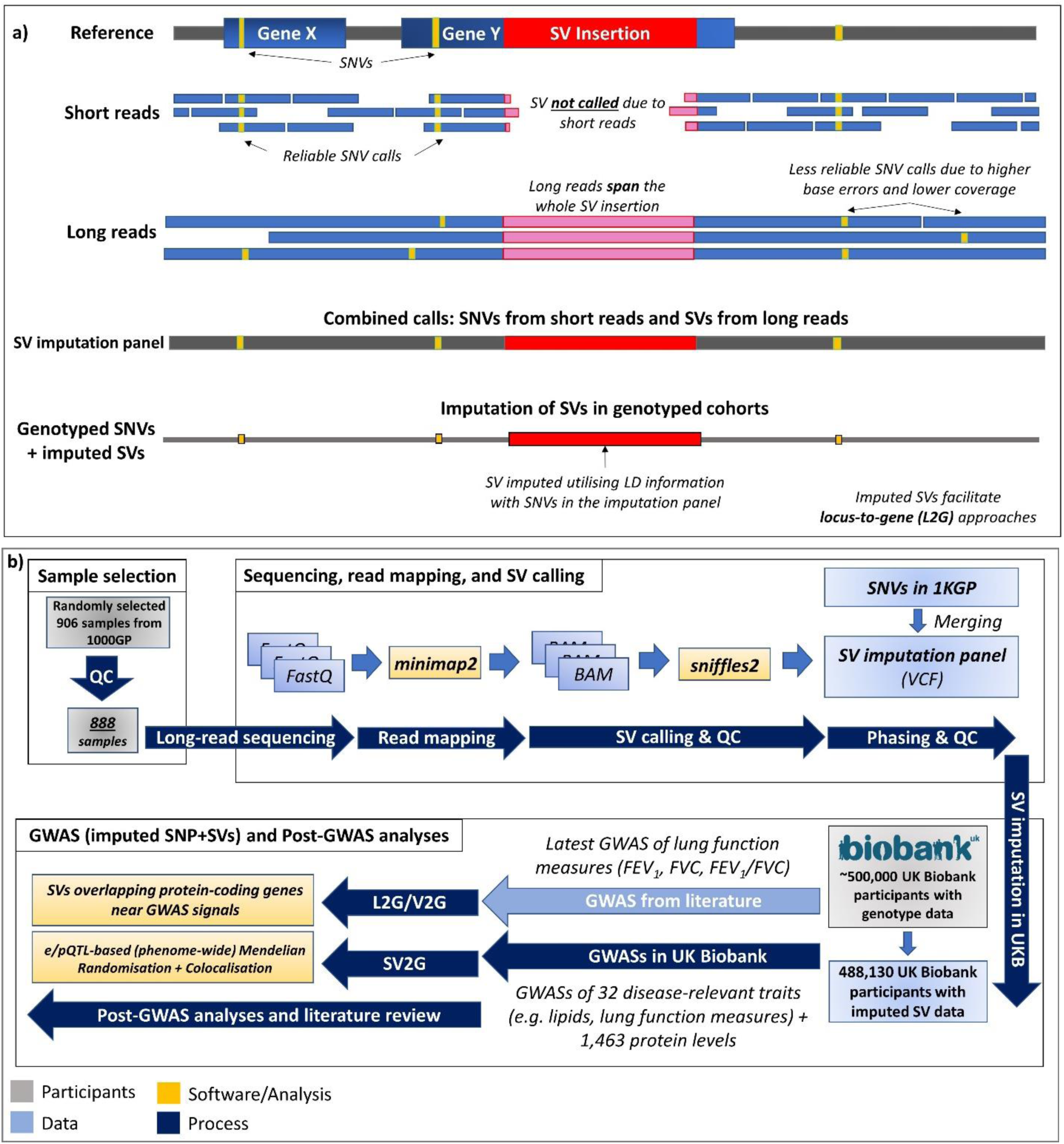
Study design and visual abstract of purpose and benefits of long-read sequencing and a multi-ancestry imputation panel. a) Long-reads that span large SVs enable identification of novel SVs and improve calling of known SVs, which better inform locus-to-gene (L2G) approaches. b) Study design including the sample selection, SV calling, SV imputation in UK Biobank, GWAS, and post-GWAS stages (not exhaustive). For the GWASs in UK Biobank, separate analyses using the same phenotype definitions were carried out using (i) the imputed SNVs, and (ii) imputed SVs (i.e., SV-WAS). The two GWAS summary statistics were then combined for the relevant post-GWAS analyses. L2G/V2G: locus-to-gene/variant-to-gene (carried out on top SNVs identified by Shrine et al., which have an SV deletion within 500 kb); SV2G: SV-to-gene (carried out **only** on SV deletions that were the most significant variants in the associated loci in the GWASs we carried out in UK Biobank, which also overlap with protein-coding genes); 1KGP: 1000 Genomes Project (Phase 1); UKB: UK Biobank.

## Results

### Structural variant calling and benchmarking

We performed long-read whole-genome sequencing of 906 individuals sampled from the 1000 Genomes Project, with a median read length of ∼6.2 kbp (**Supplementary Table 1, Supplementary Figure 1**) and 15x median coverage (alignment with minimap2; **Supplementary Table 2, Supplementary Figure 2**). The 888 samples passing quality control (QC) included 164 Europeans, 144 (admixed) Americans, 168 East Asians, 171 South Asians, and 241 Africans (**Figures 2a** and **2b; Supplementary Table 3**).

**Figure 2.**
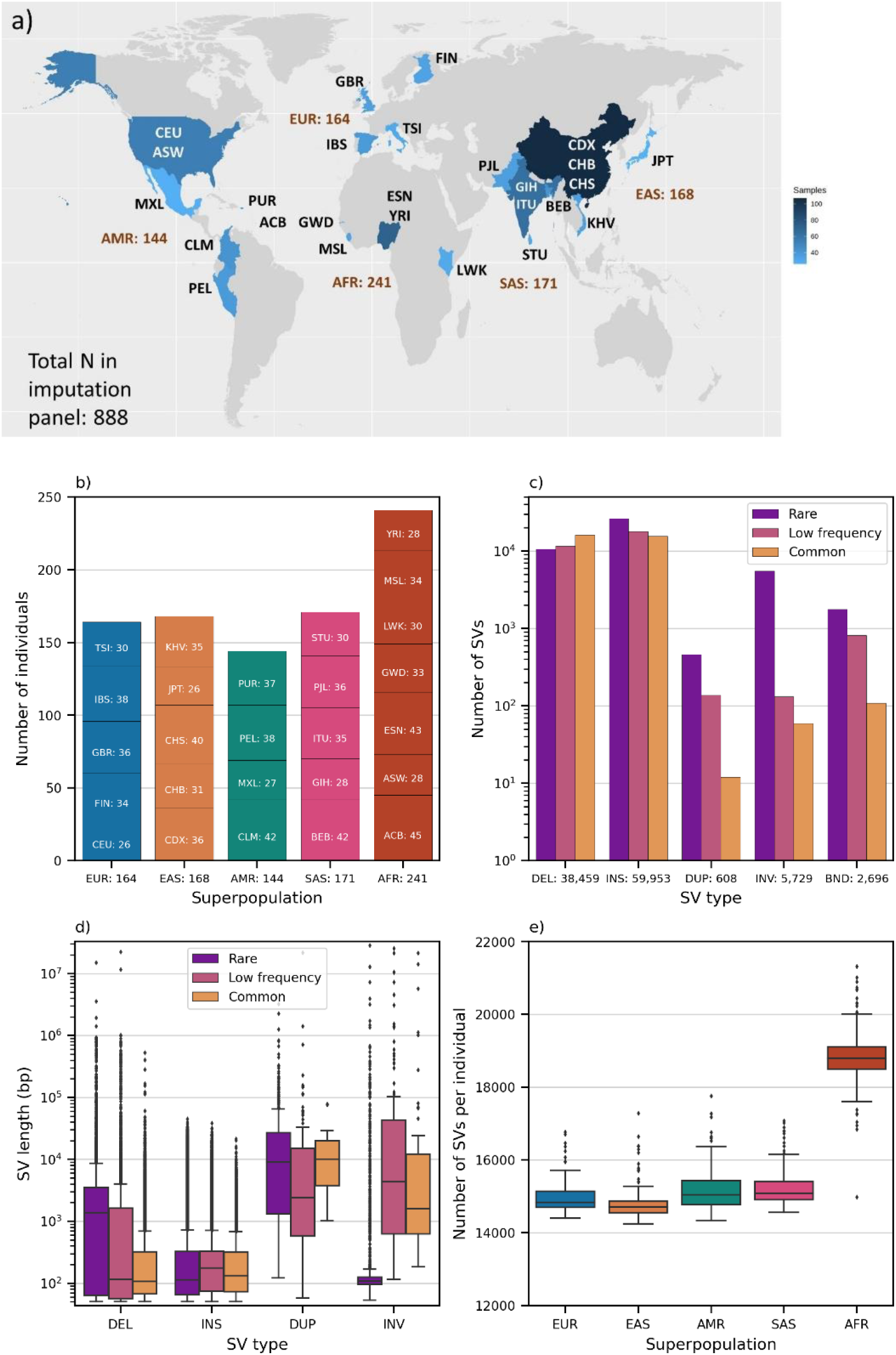
Characterisation of the quality-controlled imputation panel. (N=888 samples and n=107,445 SVs). **a)** Map of the 888 samples from the 1000 Genomes project, mapping the samples to countries and not indicating detailed geographical origins of populations. AMR: Admixed American, AFR: African, EUR: White European, EAS: East Asian, SAS: South Asian ancestries. **b)** Sample counts by superpopulation and population codes (population code description in **Supplementary Table 3**), using abbreviations from 1000 Genomes Project. **c)** Number of SVs by variant type (DEL: deletion, INS: insertion, DUP: duplication, INV: inversion, BND: break-end) and frequency class (common: 0.05<MAF≤0.5, low frequency: 0.005<MAF≤0.05, rare: 0<MAF≤0.005), with total SV counts by SV type. **d)** SV size distributions by SV type and frequency class, excluding break-ends, which do not have a size. **e)** Number of minor SV alleles per individual by superpopulation in the imputation panel.

Joint (multi-sample) calling was performed using Sniffles2^9^. To assess the quality of our SV calling, we benchmarked data generated from the 1000 Genomes project participant NA12878 (also known as HG001). This individual was analysed by us, the Genome in a Bottle (GIAB) initiative using high-coverage PacBio HIFI long-read sequencing^10^, and by Byrska-Bishop *et al.*^11^ via high-coverage short-read Illumina sequencing (referred to as the *NYGC* dataset).

We compared our raw SV calls for NA12878 to the GIAB calls of the same individual using the Truvari bench tool^12^. In a whole-genome comparison, we observed a precision of 71.8%, a recall of 76.3%, and an F1 score of 74.0%. In a second benchmark excluding tandem repeat regions longer than 200 bases, we achieved a high precision of 90.4%, a recall of 91.5%, and an F1 score of 91.0% (see **Methods** for further details). Comparing our NA12878 SV calls to the NYGC dataset, we observed a precision of 15.9% and a recall of 85.4%. This indicates that our long-read approach detected most of the SVs identified by NYGC short-read sequencing but also discovered many additional SVs. The majority (67.5%) of SVs not included in the NYGC data were present in the GIAB call set, strongly supporting the validity of our SV calls. Similarly high recall and low precision rates compared to NYGC were observed for other individuals (**Methods**, **Supplementary Figure 3**).

### SV characterisation

We used the following criteria for selecting 107,445 SVs for inclusion in the imputation panel: length between 50 bp and 30 Mbp, missingness rate <20%, and being present in at least 2 out of the 888 individuals (**Methods**, **Supplementary Table 4, Supplementary Figures 4 and 5**). Based on the benchmarking of our SV calls against the NYGC call set, we annotated SVs in the panel with three categories: “novel”, SVs not detected in the NYGC dataset (75,192 or 70%); “known”, SVs previously identified in at least one individual (29,119 or 27.1%); and “not compared”, SVs ignored by the Truvari bench tool (3,134 or 2.9%) (see **Methods**).

The GIAB consortium described which genomic regions confidently produce reliable genotypes^13^, and 41.9% of our SVs mapped to high-confidence (‘confident’ henceforth) regions. The most common SV types were insertions (55.8%), followed by deletions (35.8%), inversions (5.3%), break-ends (typically unresolved SVs; 2.5%), and duplications (<0.06%) **(Figure 2c)**. Most SVs were rare (minor allele frequency (MAF) <0.005: 41.8%), the remainder being low-frequency (MAF 0.005-0.05: 28.7%) and common (MAF >0.05: 29.6%) variants, with the latter being enriched for insertions and deletions. Note that the panel contains a larger proportion of rare SNVs than rare SVs because a) rare SVs are more difficult to detect reliably compared to rare SNVs and b) the SNVs were originally called in the larger, full 1000 Genomes cohort.

Sizes of most SV deletions and insertions were between 50 and 1000 bp (**Figure 2d**). Duplications and inversions predominantly ranged from 1 to 30 kbp. A considerable number of SVs extended up to 1 Mbp and beyond. However, SVs that are longer than 1 Mbp are likely to be artefacts of the SV calling process (see **Methods**).

Of the 107,445 SVs, 4,406 SVs were predicted using SnpEff^14^ to have high functional impact. Of these, 1,198 were predicted to cause frameshifts, and 581 the complete loss of exons. Based on GWAS Catalog^15^, 2,465 SVs overlapped positions of variants with known GWAS associations (**Supplementary Table 5**).

On average, individuals carried 16,065 SVs, with individuals of African ancestry exhibiting the highest (mean: 18,822 SVs), and East Asian-ancestry individuals exhibiting the lowest (14,729) SV diversity (**Figure 2e, Supplementary Table 6**)^3,16^. This observation was most pronounced for insertions and deletions, which are more frequent and can be called with high confidence. Only 36.6% (39,273) of SVs were shared across all superpopulations with individuals of African ancestry exhibiting the most SVs not shared with other populations (13,153) and (Admixed) Americans the least (841) (**Supplementary Table 7**).

### Generation of SV imputation panel and application to UK Biobank

To enable the imputation of SVs in genotyped studies, we constructed a haplotype reference panel consisting of 888 samples. To this end, we merged the 107,445 SVs generated in the present study with the ∼45M short variants identified from short-read sequencing data of the 1000 Genomes Project Phase 3 release^11^ (see **Methods**).

We assessed the suitability of the panel for SV imputation into UKB by performing leave-one-out imputation for all individuals in the panel. Here, we specifically emulated the process of SV imputation in UKB by using all genotyped UKB SNVs present in the reference panel (∼702K SNVs) as the basis for imputation. To facilitate benchmarking, we not only imputed SVs but also ∼58K randomly selected SNVs from the panel. We calculated the per-variant non-reference genotype concordance^17^ and the imputation quality metric *r^2^_imp_* (see **Figure 3b**; **Methods**; **Supplementary Table 9, Supplementary Figure 6**). Both metrics varied based on MAF, GIAB region type, and variant type. Generally, rare SVs showed poorer imputation quality than common insertions and deletions, which produced high-quality imputation metrics. The mean non-reference concordance for common insertions and deletions was 0.696 and 0.693, respectively (both concordances were 0.846 in confident regions), with mean *r^2^_imp_* =0.718 and 0.721 (0.858 and 0.852 in confident regions, see **Supplementary Table 9 and Figure 3b**). These metrics demonstrate sufficient imputation quality for conducting GWAS of common variants. As anticipated, the imputation quality of SVs was lower than that of imputed SNVs across GIAB region types and MAF classes, due to the greater difficulty in reliably calling SVs; however, this difference was not substantial in confident regions (**Figure 3b**). Note that, for our SV association analyses (see below), we only evaluated results for high-quality SVs.

**Figure 3.**
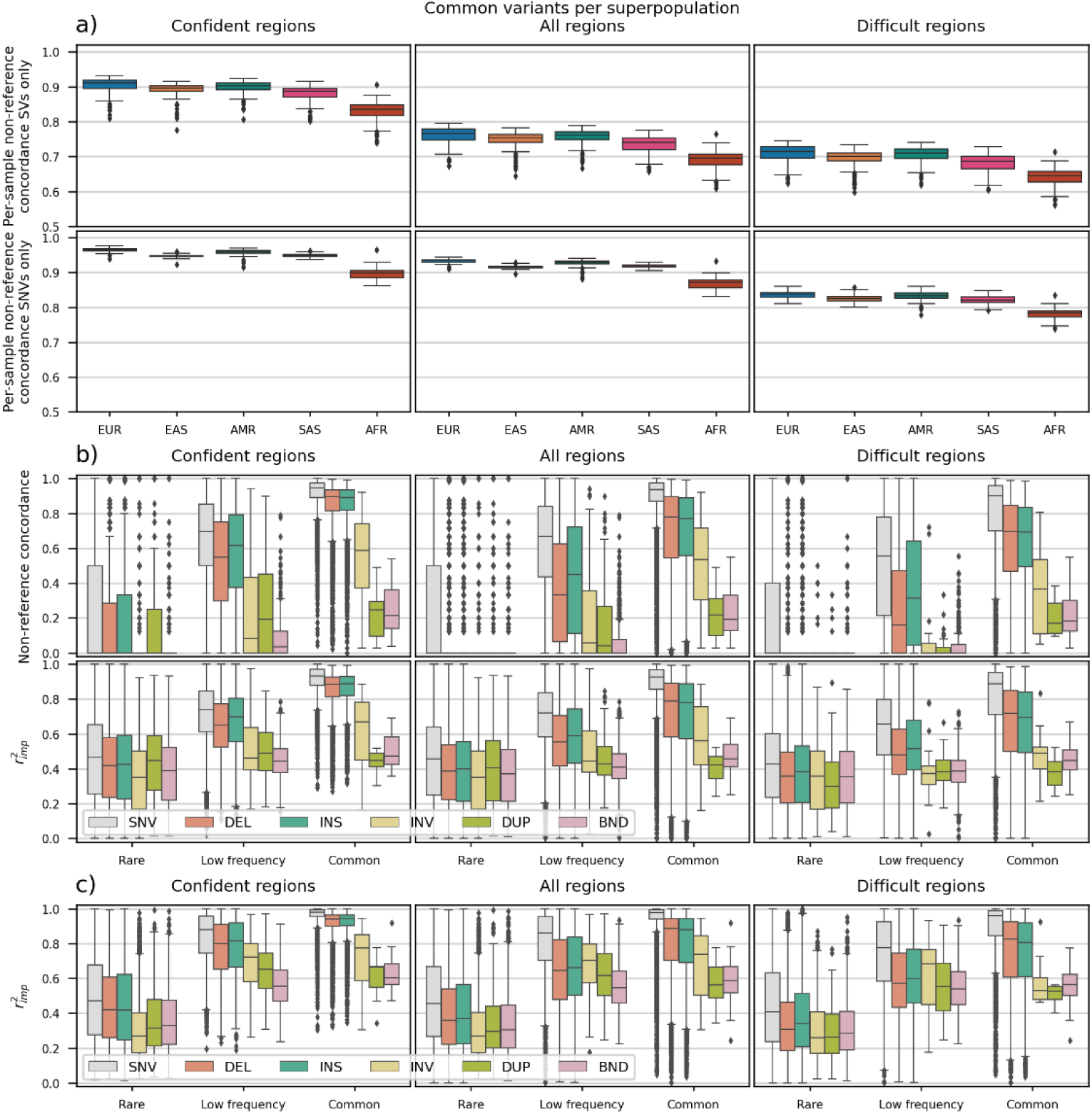
Evaluation of imputation quality: a) Leave-one-out imputation: sample-wise non-reference concordance stratified by superpopulation and GIAB region type, based on imputed common SVs (top) and imputed common SNVs (bottom). b) Leave-one-out imputation: variant-wise non-reference concordance (top) and *r^2^_imp_* score (bottom) stratified by variant type, MAF class, and GIAB region type. c) Imputation into UK Biobank: *r^2^_imp_* score by variant type and MAF class, for confident regions, all regions, and difficult regions.

We also computed the per-individual non-reference genotype concordance for each superpopulation, based on common SVs and SNVs stratified by GIAB region type (**Figure 3a**; **Supplementary Table 8**). African-ancestry individuals, being the most diverse, had a mean non-reference concordance of 0.690 based on all common SVs, which was the lowest amongst all superpopulations. By comparison, individuals of European ancestry showed a mean non-reference concordance of 0.760 for common SVs. Overall, we did not observe significant outliers in the per-individual concordance values, indicating that there were no issues with sequencing and data processing of the samples. In conclusion, the SV reference panel offers a robust foundation for imputing SVs, particularly common insertions and deletions, into UKB and for performing subsequent GWAS.

We used our imputation panel to impute SVs into the genomes of 488,130 UKB participants (see **Methods**). Imputation quality in UKB mirrored our observations from the leave-one-out validation in the 1000 Genomes study: *r^2^_imp_* was higher for common variants and in high-confidence regions. For example, common deletions and insertions in high-confidence regions showed mean *r^2^_imp_* =0.915, compared to 0.961 for common SNVs in the same regions (**Supplementary Tables 10** and **11**; **Figure 3c**). Notably, UKB predominantly consists of European-ancestry individuals, for which we observed higher average imputation qualities in the 1000 Genomes leave-one-out analyses.

Finally, as a proof of principle, we manually compared one deletion (Sniffles2.DEL.3639MF) imputed from the genotype data to the recent UKB short-read based whole-genome sequencing (WGS) data^18^. To this end, we analysed the read coverage from *vcf* files of the respective genomic region to determine chromosomes carrying the deletion allele (**Methods** and **Supplementary Figure 7**). Thus, we could group individuals based on their ‘Sniffles2.DEL.3639MF’ deletion genotype and compare them to the imputed genotypes (**Supplementary Figure 8)**. We found a high concordance rate of 98.7% between the WGS and imputed genotypes, exceeding the accuracy estimations from the leave-one-out analyses. We chose this specific deletion for the proof-of-principle analysis because it a) has an interesting disease association and is thus relevant (see Figure 4) and b) our calling approach for SVs from WGS data only works for deletions in non-repetitive regions (see **Methods**).

**Figure 4.**
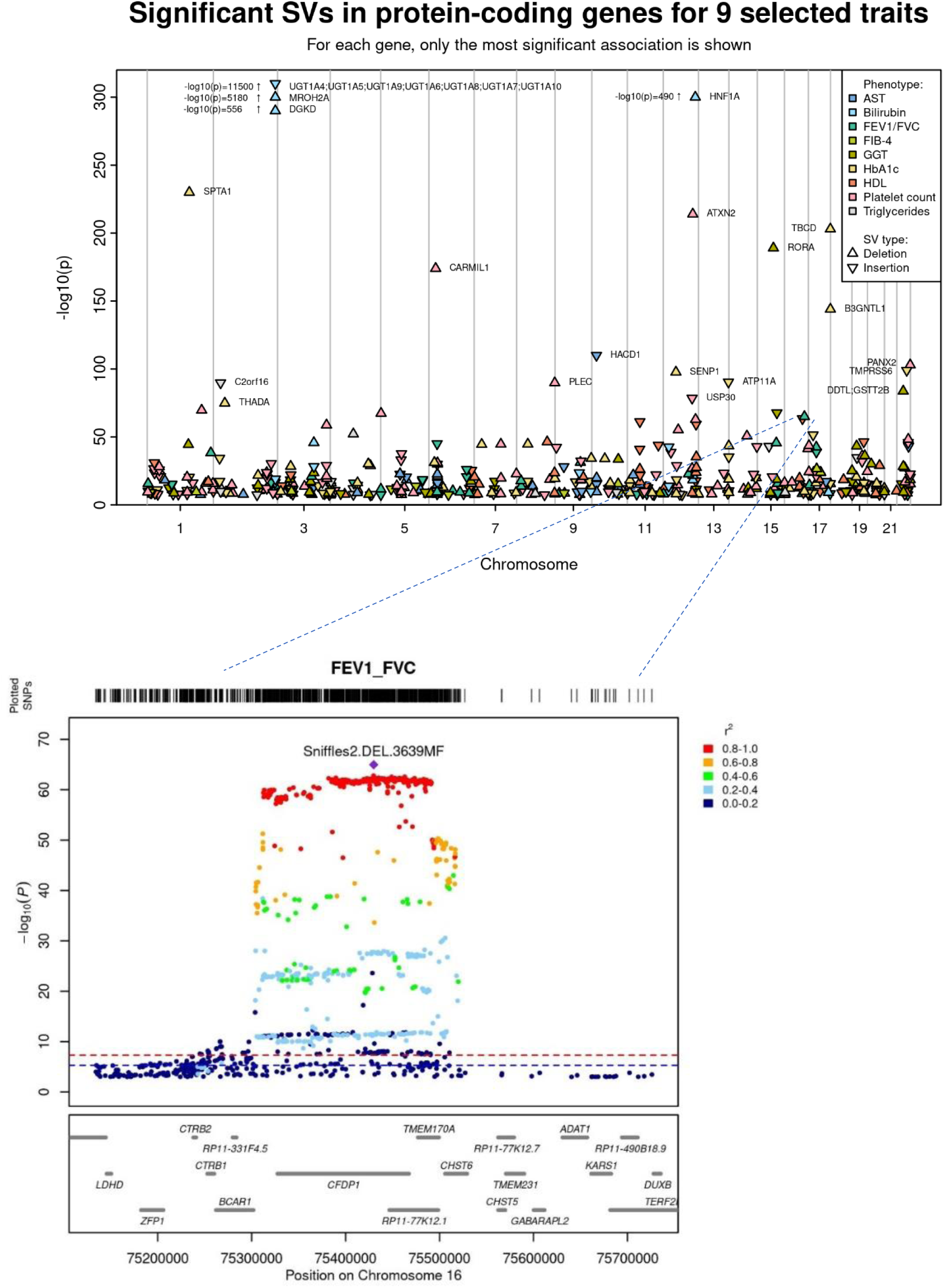
**a)** Manhattan plot of 9 selected traits, with only GW-significant SVs overlapping with protein-coding genes shown. Phenotype definitions and full list of GW-significant SVs are available in **Supplementary Tables 12-13** and **14**, respectively. **b)** Region plot showing the FEV_1_/FVC association of Sniffles2.DEL.3639MF (841-base SV deletion in an intron of *CFDP1*) and nearby SNVs in a GWAS of UK Biobank participants. LD information between the variants was calculated using the same population utilised in the UKB GWASs. Additional details on the SV can be found in **Supplementary Table 5**.

### Proof-of-principle SV imputation and genome-wide association studies in UK Biobank

To demonstrate the added value of the imputation panel, we selected 19 exemplary continuous traits and 13 binary traits available in UKB relevant to respiratory (n=6 traits), metabolic (n=16), and liver (n=10) diseases (**Supplementary Tables 12** and **13**). On these phenotypes, we performed GWASs of the imputed SVs (SV-WAS) in up to 453,754 European UKB participants^19^ (**Supplementary Figure 9**).

Filtering variants by imputation metric INFO>0.7 and MAF>0.01, 3,858 SV associations (in 1,898 unique SVs, 1,258 of which were in the *novel* category) passed the established genome-wide significance threshold of *p*<5×10^−8^ in any phenotype (**Supplementary Table 14**). For an in-depth assessment of potential causal genes, we selected all significantly associated SVs from these SV-WASs that overlapped functional protein-coding genes (some SVs spanned several genes), thereby prioritising 689 unique genes (**Figure 4a**; **Supplementary Table 14**; **Methods**). We also analysed how SVs influence the levels of 1,463 proteins in blood plasma. Here, we identified 10,518 significant (*p*<5×10^−8^/1463=3.4×10^−11^, INFO>0.7, MAF>0.01) SV-based pQTLs (3,723 unique SVs, 2,495 of which were novel) for 1,101 proteins (**Supplementary Table 15**), including 84 (excluding the major histocompatibility complex region, *i.e.*, *6p21*) that were conditionally independent of nearby SNVs (**Methods**; **Supplementary Table 16**).

### Systematic analysis of SV information in identifying novel associations and causal genes

To follow up on the significant SV associations identified in the UKB SV-WAS and pQTL analyses, we examined whether these SV associations provide added value compared to analysing SNVs alone. To this end, we carried out association analyses on imputed UKB SNVs mapping to each locus containing significant SVs, using the same sample inclusion criteria, phenotype transformations, and covariates as in the SV analyses. To identify whether SVs showed more robust evidence for an association, *i.e.*, lower *p*-values, at the SNV-based GWAS- and pQTL-associated loci, we combined the SNV and SV association results. At 55 genome-wide significant GWAS loci, an SV showed the lowest *p*-value (**Supplementary Table 17**). Conditional analyses revealed that SVs constituted a secondary, independent signal at 23 further loci; 38 of these 78 SVs overlapped with protein-coding genes.

We then systematically assessed the contribution of SV associations to the prioritisation of causal genes by comparing genes implicated by SVs to the putatively causal genes reported by the latest GWAS of lung function measures published by Shrine *et al.*^20^. The authors used several post-GWAS approaches (e.g., nearest gene, e/pQTLs, polygenic priority score (PoPS, a tool for gene prioritisation^21^), and rare respiratory disease causal genes) to map associated loci to genes, thereby prioritising on average ∼3 genes per locus. We examined the reported autosomal loci associated with ≥1 of the three lung function phenotypes (*i.e.*, FEV_1_, FVC, or FEV_1_/FVC – the latter a clinically important lung function measure utilised in diagnosing chronic obstructive pulmonary disease). Seventy of these reported loci harboured SVs (*i.e.*, an SV mapped within ±500kb of the top SNV) that were significantly associated with the same primary lung function trait in our UKB-based SV-WASs (**Supplementary Table 18**). At 55 of these 70 loci, the gene implicated by our SV analyses was also implicated by ≥1 of the post-GWAS methods utilised by Shrine *et al.* – strengthening the evidence for causality for the respective genes. In the remaining 15 loci where SV-implicated genes had not been prioritised yet, SVs pointed to genes very likely involved in lung health via influencing smoking behaviour (e.g., *SLC1A2* - highly expressed in the brain; **Supplementary Figure 10**) or other lung-disease causal mechanisms such as abnormal respiratory ciliary function^22^ (e.g., *DNAH12* and *DYNLRB1*) and promotion of inflammation^23^ (e.g., *PRDX1*).

### Added value of SV information to gene prioritisation: examples from GWASs of lung function measures

In addition to the identification of the above-mentioned genes, the added value of SV information for identifying the causal gene, *i.e.*, improving locus-to-gene (L2G) prioritisation pipelines at the loci identified by Shrine *et al.*^20^, can be demonstrated using four representative examples:

In our association analyses of quantitative lung function measures in UKB, SVs constituted the conditionally independent variant in primary or secondary signals of 14 loci, thereby facilitating identification of the respective causal genes (**Supplementary Table 17**). One of these FEV_1_/FVC-associated loci contained an 841-base SV deletion in an intron of *CFDP1* (Sniffles2.DEL.3639MF; p=1.1×10^−65^; MAF=0.413; **Figure 4b**). We confirmed this deletion using WGS data. Shrine *et al.* prioritised three potentially causal genes at this locus (including *CTRB1* and *BCAR1*), with *CFDP1* only being implicated by its proximity to the top SNV (rs11864587) without any functional or regulatory support. Follow-up analyses on this gene, including phenome-wide cis-eQTL-based Mendelian Randomisation (MR) and colocalisation analyses (not carried out by Shrine *et al.*) provided strong evidence (min MR *p*=1.1×10^−22^; colocalisation posterior probability (PP) >90%) for putatively causally linking *CFDP1* expression in various (bulk GTEx) tissues including fibroblasts – a cell type relevant for respiratory disease – to a lower FEV_1_/FVC measure (**Supplementary Figure 11**; **Supplementary Table 19**). *CFDP1* and *BCAR1* protein expression levels were not assayed by the UKB-PPP^24^, thus information from SV-based pQTLs could not be utilised to distinguish between the genes.

At a second FEV_1_/FVC-associated locus (top-associated SNV: rs947350), Shrine *et al.* prioritised ten genes, including *MEGF6* with suggestive evidence from rare coding variants. In our study, an FEV_1_/FVC-associated SV deletion (Sniffles2.DEL.6E8M0; p=2.1×10^−16^; MAF=0.069) mapped only to *MEGF6*. Similar to the *CFDP1* example, phenome-wide MR and colocalisation analyses provided strong evidence (min MR *p*=6.3×10^−7^; colocalisation PP>97%) putatively causally linking *MEGF6* expression in various (bulk GTEx) tissues including skeletal muscle and heart tissue (a likely respiratory disease-relevant cell type as a smooth muscle-containing tissue) to a lower FEV_1_/FVC measure (**Supplementary Figure 12**; **Supplementary Table 20**).

Third, *AAGAB* was solely implicated by two different FVC-associated SV deletions (Sniffles2.DEL.268AME; p=1.6×10^−12^; MAF=0.231; and Sniffles2.DEL.2689ME; p=2.6×10^−12^; MAF=0.233) but not by any of the post-GWAS methods utilised by Shrine *et al*. Similar to the above examples, phenome-wide MR and colocalisation analyses provided strong evidence (min MR *p*=1.7×10^−15^; colocalisation PP>90%) putatively causally linking *AAGAB* expression in various (bulk GTEx) tissues, including lung tissue, directly to lung function measures (**Supplementary Figure 13**; **Supplementary Table 21**).

As a final example, Shrine *et al.* identified rs7108992 to be associated with FVC and FEV_1_. They prioritised two genes, *ETS1* (by proximity) and *FLI1* (via PoPS). In our analysis, we identified an FVC-associated SV deletion only mapping to *FLI1* (Sniffles2.DEL.5C9EMA; p=2.0×10^−30^; MAF=0.322). Phenome-wide MR and colocalisation analyses strongly linked *FLI1* expression in lung and smooth muscle-containing tissues to lung function measures (MR *p*=3.9×10^−4^; colocalisation PP=96%), pulmonary heart disease risk (MR *p*=8.8×10^−7^; colocalisation PP=80%), *FGF10*^25^ (MR *p*=7.5×10^−5^; colocalisation PP=89%), and *LRP1* protein levels^26^ (MR *p*=2.8×10^−5^; colocalisation PP=87%) – known factors contributing to respiratory diseases^25^ (**Supplementary Figure 14**).

Taken together, these examples illustrate the potential added value of SV information for the identification of novel gene-disease associations, and for improving gene prioritisation pipelines applied to GWAS summary statistics (e.g., the composite locus-to-gene (L2G) score calculated in Open Targets Genetics^27^).

## Discussion

Long-read sequencing holds the promise of conducting reliable association studies of SVs in large cohorts, but its widespread adoption is impeded by its significant cost. For example, comprehensive long-read sequencing of all UKB participants would cost approximately 0.5 billion USD, based on an estimate of 1,000 USD per whole genome. It was recently suggested to reduce costs by sequencing only a limited number of SVs^28^. By contrast, our SV imputation approach allows for analyses of a comprehensive, genome-wide SV panel without additional sequencing costs. Therefore, use of an SV imputation panel constitutes a practical and cost-effective solution for the robust analysis of common SVs. The multi-ancestry imputation panel applied in the present study to Europeans from UKB can also be used to impute SVs in diverse ancestries, e.g., from BioBank Japan^29^, China Kadoorie Biobank^30^, or the Singapore Precision Medicine Programme^31^.

In this context, we observed that the number of SVs detected per individual differed between superpopulations. We identified the highest average number of SVs in individuals of African descent and a slightly lower average in East Asians, compared to the other superpopulations. While we included a higher number of African ancestry individuals, the number of East Asian individuals included in our reference panel was comparable to the number of individuals from other, non-African ancestries. In fact, it was even larger than the number of European ancestry individuals (AFR n=241, SAS n=171; EAS n=168; EUR n=164; AMR n=144). Therefore, we do not expect a major bias from underrepresentation of any superpopulation in the training dataset. It is well established that African ancestry is more diverse than is the case for other superpopulations^32,33^ and previous studies indicate that East Asian populations tend to exhibit slightly lower genetic diversity compared to European populations^34^, which is consistent with the lower observed SV counts per genome.

We benchmarked SV calls of the individual NA12878 against GIAB’s PacBio-based calls, achieving both high precision and recall, and found that our SV calls, compared to Illumina’s short-read data, detected most known but also many additional (‘novel’) SVs, with validation from GIAB’s PacBio dataset. Seventy percent of the SVs contained in our imputation panel can be considered as novel, demonstrating the value of long-read sequencing in SV identification efforts. The size of the sample used for our long-read sequencing was smaller than some of the short-read sequencing samples available. However, our analyses have demonstrated that long-read sequencing is required to detect most of the novel SVs identified in the present study.

We observed that the quality of SV calling and imputation is strongly stratified by variant type, frequency, and the complexity of genomic regions. Using genome stratification files provided by the GIAB consortium, we identified robust SV calls and showed that deletions and insertions with a MAF>0.05 were most reliable. These insights motivate future refinement of variant stratification methods.

We demonstrated the utility of our SV imputation panel by imputing SVs in UKB and conducting SV-WASs on 32 disease-relevant traits. Here, 907 SVs were significantly associated, mapping to 689 functional protein-coding genes. We also ran association analyses of ∼1.5k plasma protein levels and identified 1720 significant SVs overlapping with 1197 genes. The majority of SVs carrying a significant association with the traits or protein levels were novel, *i.e.*, not detected in 1000 Genomes short-read sequencing. Using selected examples, we highlighted the relevance of the identified potentially causal genes to respiratory diseases.

In the present proof-of-principle study, we focused on SVs overlapping with the coding sequence of genes. In future applications of our SV imputation panel, more refined gene mapping approaches could be employed, e.g., including SVs overlapping enhancer regions or epigenetic marks. Such an enhanced mapping would increase the number of identified associated genes and thus provide additional insights into regulatory mechanisms and disease biology.

Given the sample size of our imputation panel, the resource might be more useful in fine-mapping signals at known GWAS-associated loci than to identify novel disease-associated loci. Our imputation panel facilitates analyses of SVs overlapping exons, splice-sites, promoters, or enhancers of protein-coding genes at GWAS-associated loci. Therefore, it has the potential to become a routine component of post-GWAS gene prioritisation workflows. Structural variants can influence complex disease biology through either the disruption of coding sequence or an altered regulation of gene expression. Such effects may not be well captured by short variants alone. Incorporating SVs into GWAS and follow-up analyses would thus provide more accurate disease risk prediction, uncover underlying pathomechanisms by highlighting actionable pathways and targets, and support precision medicine by providing biomarkers for patient stratification. We also envisage that our SV imputation panel will enable diverse applications of integrating SVs with other *omics* data, e.g., in machine learning-based frameworks. For example, genome-wide SV-based features could be used in models for high-throughput post-GWAS gene prioritisation^27^, disease subtyping/precision medicine^35,36^, and drug response/adverse event prediction^30^.

As next steps in applying our SV panel resource, we suggest, first, to impute SVs in other international biobanks, making more SV data available to the research community. Second, to carry out GWASs utilising SVs to identify novel associations and disease-causal genes. Finally, to carry out comprehensive analyses of the effects of SV inversions (e.g., spanning transcription factor binding sites), translocations, and duplications (e.g., to analyse dosage-dependent effects of SNVs affected by the duplications) for which more research is needed. Thereby, this resource will strongly advance our knowledge of the genetic underpinnings of disease.

## Methods

### Oxford nanopore long-read sequencing

Sample sequencing was executed using Oxford Nanopore Technologies’ (ONT) PromethION P48. After DNA extraction, libraries were prepared according to ONT ligation sequencing kit SQK-LSK110 protocols with r9.4.1 flowcells (further details can be found in the Qiagen Gentra Puregene Handbook).

For base calling, we used guppy v6.0.1^37^ with the super high accuracy model (SUP). Sequencing statistics for the individual *fastq* files were generated with the NanoStat^38^ software (**Supplementary Table 1; Supplementary Figure 1**).

The data generation process for the complete long-read dataset is detailed in Schloissnig et al.^7^

### Alignment and calling

The alignment was performed against the GRCh38 assembly using minimap2 v2.24^39^ and recommended default parameters for Oxford Nanopore long reads (“-ax map-ont”).

Alignment metrics were computed with Picard’s^40^ CollectWgsMetrics tool, with modified parameters to adapt for Oxford Nanopore reads and our analysis setup. Specifically, Picard was instructed to count unpaired reads (“--COUNT_UNPAIRED true”), all base qualities (“--MINIMUM_BASE_QUALITY 0”), and only coverages at sites in the reference on autosomal chromosomes (interval file via “--INTERVALS”). For descriptive statistics see **Supplementary Table 2**.

Joint variant calling was conducted with Sniffles2^9^ v2.0.7, supplemented by tandem repeat annotations to improve variant calls in these regions, using the default annotation file provided by the tool’s authors (“--tandem-repeats human_GRCh38_no_alt_analysis_set.trf.bed”).

### Sample identity and quality verification

We performed extensive sample identity verification. We generated long-read ONT sequencing data for 906 DNA samples from the 1000 Genomes project. To confirm the identity of 906 ONT-sequenced individuals, we used the aligned reads to call genotypes for each DNA sample at approximately 12,000 common SNVs (with AF between 0.45 and 0.55) across the genome. We ran ‘bcftools mpileup’ on each sample bam file with the following settings:

bcftools mpileup $bam -f $hg38_fasta --skip-indels -a FORMAT/AD -B -R SNV_coordinates.txt -X ont The file ‘SNV_coordinates.txt’ contained the genome coordinates of the SNVs.

Next, we extracted the genotypes of the same SNVs from 3,202 individuals of the 1000 Genomes Project Phase 3 release *vcf* files using ‘bcftools view’. For each ONT-sequenced DNA sample, we then generated a vector of read ratios, where each vector element was a number between 0 and 1 – the number of reads carrying the non-reference base at the SNV site divided by the total number of reads. We converted the 1000 Genomes Project Phase 3 genotypes to comparable numerical vectors by substituting homozygous reference, heterozygous, and homozygous non-reference genotypes with 0, 0.5, and 1, respectively.

To compare the SNV genotypes of ONT-sequenced and 1000 Genomes phase 3 individuals, we calculated the pair-wise distance between ONT and 1000 Genomes phase 3 genotypes. As a distance measure, we used the square of the Euclidean distance divided by the number of SNPs. Since the AFs of the selected SNVs were close to 0.5, the distance measure between unrelated samples is close to 1, around 0 for identical samples, and around 0.5 between first-degree relatives. Moreover, these distances also allowed us to identify contaminated sequencing runs within the ONT data. In case of contamination, the observed distance between DNA samples from supposedly unrelated individuals ranges between 0 and 1, depending on the extent of the contamination.

### Sample exclusions and corrections

In total, 18 sequenced DNA samples were removed and the labels of three were corrected: After careful analysis, seven samples were found to be contaminated, and therefore removed from the panel: HG02813 (contaminated with HG02807), HG02870 (contaminated with HG02813), HG02888 (contaminated with HG02870), HG02890 (contaminated with HG02888), HG03778 (contaminated with HG03793), and HG00138 (contaminated with HG0017), and HG02895 (contaminated with HG02890). Furthermore, we found that the DNA sample labelled as HG02807 belonged to individual HG02895 and corrected the label accordingly. Individuals HG01951 and HG01983 were swapped during sequencing and we swapped the labels back.

Seven samples were removed because they were descendants of other individuals in the panel, namely: NA19828, HG03804, HG00420, HG01258, NA19129, NA12877, and NA12878. Note that the SV calls of NA12878 were still used for SV calling benchmarking, see below. Two additional individuals were removed due to sequencing problems, including HG00128, which had 50.1% duplications, and NA18966, which had 34.5% duplications in one of the two *fastq* files available for the sample.

In addition, individual NA15728 was found to be not from the 1000 Genomes dataset and was therefore removed from the panel. Finally, individual HG03127 was removed from the panel due to low quality and low performance metrics in the preliminary leave-one-out analysis. Thus, 18 DNA samples were removed, resulting in 888 individuals included in the imputation reference panel.

### Benchmarking SV calls

We generated a benchmark of our SV calls for the NA12878 individual (also known as HG001) against the Genome in a Bottle (GIAB) set of SV calls for the same individual based on high coverage 15kb and 20kb PacBio HIFI sequencing (see https://ftp-trace.ncbi.nlm.nih.gov/giab/ftp/data/NA12878/ analysis/PacBio_CCS_15kb_20kb_chemistry2_042021/).

We ran the Truvari bench v4.2.2 tool, with settings ‘--passonly --pctseq 0 --pctsize 0.5 --refdist 500 --dup-to-ins’ to compare our SV calls for NA12878 (the comparison set) to the SV calls included in a file HG001.GRCh38.pbsv.vcf.gz from the link above (the base set). The true positive set accounted for 17,571 variants, while the false positive and false negative sets included 6,899 and 5,464 SVs, respectively. This resulted in a precision of 71.8%, a recall of 76.3%, and an F1 score of 74.0%.

We also generated the benchmark excluding tandem repeat regions, which are known to be challenging for accurate SV calling, because the same SV can be placed in different places in the tandem repeat. The benchmark achieved a very high precision (90.4%), recall (91.5%), and F1 score (91.0%), with true positive, false positive, false negative sets comprising 8486, 897, 789 SVs, respectively. The definition of tandem repeat regions that was used can be found in https://raw.githubusercontent.com/PacificBiosciences/pbsv/master/annotations/human_GRCh38_no_alt_analysis_set.trf.bed. We only excluded tandem repeats longer than 200 bases, there were 172,364 such repeats. The non-tandem repeat regions were included in the Truvari bench run by adding the ‘--includebed human_GRCh38_no_alt_no_trf200.bed.gz --extend 500’ parameters. The *bed* file used here was generated with the command ‘bedtools complement’ for tandem repeats longer than 200 bases.

We also compared our SV calls of NA12878 against the NYGC SV call set based on short-read high-coverage Illumina sequencing from Byrska-Bishop *et al*.^11^ To obtain the published call set for comparison, we downloaded the *VCF* files containing the phased SNVs, InDels, and SVs (https://ftp.1000genomes.ebi.ac.uk/vol1/ftp/data_collections/1000G_2504_high_coverage/working/20220422_3202_phased_SNV_INDEL_SV/), extracted SVs (variant IDs starting with ‘HGSV’) present in the NA12878 sample, and ran ‘Truvari bench’ with the same settings as above, now using the short-read-based SV callset as the base set.

The benchmark identified 3,675 true positive SVs, 19,446 false positives, and 630 false negatives, resulting in a precision of 15.9%, a very high recall of 85.4% and an F1 score of 26.8%. This indicates that we successfully detected most SVs identified by short-read sequencing and also identified many additional SVs. Notably, a large proportion (13,127 out of 19,446, or 67.5%) of these additional SVs were also present in the PacBio-based GIAB call set for NA12878 described above, strongly supporting the validity of our calls.

We used the same procedure to benchmark our SV calls for all the 888 samples in the panel, comparing each sample individually with the NYGC SV calls in the same individual. The recall and precision numbers were similar to that of NA12878 (**Supplementary Figure 3A**), with recall rates around 83-87% and precision around 16-17% for the majority of non-African ancestry samples. For African ancestry individuals, recall was around 80% and precision around 18-19%. Excluding SVs in tandem repeats longer than 200 bases from the comparison increased the recall to around 90% (**Supplementary Figure 3B**) and doubled the precision to around 34% for non-African ancestry samples. The difference in the recall and precision rates between African ancestry and other individuals (the two clusters on **Supplementary Figures 3A** and **B**) can be attributed to the higher number of SVs per individual in case of African ancestry, as shown in **Figure 2e**.

Following the SV comparison analysis described above, we annotated each SV in the panel as one of three categories: “novel” – not present in the true positive set for any of the 888 individual-level comparisons with the NYGC-generated SVs; “not compared” – SVs that are ignored by the Truvari bench tool, *i.e.*, break-end (BND)-type SVs or SVs longer than the default size cut-off of 50 kbp; or “known” – SVs classified as true positives in the benchmark for at least one individual.

### Individual-level genotype missingness rates

The individual missingness rates exhibited substantial differences mainly due to variation in coverage, with over 10% missing data in 65 samples (**Supplementary Figure 4**). In this context, we adopted a lenient QC approach for several reasons: Primarily, our aim was to enhance sensitivity and we observed a significant stratification of missingness rates by variant type and region *difficulty* (**Supplementary Figure 5**). For instance, the sample-wise median missingness rates for all, confident, and difficult regions were 0.42%, 0.11%, and 0.63%, respectively. In contrast, the average missingness rates for the same regions were 2.60%, 2.42%, and 2.74%, respectively.

By adopting this approach, we were able to leverage the data from samples with higher missingness rates in regions where successful genotyping was achieved. Furthermore, we could filter for SVs with elevated missingness rates in our final association results, thereby optimising the use of available data.

### Variant-level missingness rates

Overall, raw genotype calls exhibited elevated missingness rates. However, these rates were noticeably stratified by SV type and region type (*i.e.*, difficult and confident regions, see **Supplementary Figure 5**, **Supplementary Table 22**). Deletions exhibited the highest mean missingness rate at 0.166, closely followed by insertions with a mean missingness rate of 0.086. In contrast, the missingness rates within confident regions were considerably lower, registering at only 0.023 and 0.025 for deletions and insertions, respectively. Other SV types within confident regions also maintained low missingness rates, ranging from 0.027 to 0.029.

We removed SVs with a genotype missingness rate >0.2, thereby excluding 15,830 SVs. Thereafter, the mean missingness rates were uniform across SV types: they ranged from 0.023 to 0.028 in confident regions and had slightly higher values of 0.026 to 0.033 for difficult regions. In both region types, deletions showed the highest and break-ends the lowest call rates.

These results demonstrate that although genotype missingness rates can be elevated in SV calls, problematic calls are predominantly localised within genome sections that are challenging to sequence. Therefore, high missingness rates can be effectively managed by stratifying SVs by region type and excluding genotypes with very low call rates.

### False-positive calls of very large SVs generated by Sniffles2

It is known that Sniffles2 (as of version 2.0.7) generates false-positive calls for inversions (INV), duplications (DUP), and deletions (DEL), with apparent lengths up to the size of chromosomes. This issue has been previously reported on the Sniffles2 GitHub issue tracker^41,42^. Examples in our dataset include a 149 Mbp duplication on chromosome 4, a 131 Mbp inversion on chromosome 1, and a 64.5 Mbp deletion on chromosome 3. These unreasonably large (and hence false-positive) SV calls are a consequence of the internal logic of Sniffles2: Sniffles2 cannot handle edge cases with complex rearrangements, e.g. cut-and-paste insertions with simultaneous inversion of the insertion sequence. It is currently unknown whether insertions (INS) or break-ends (BND) can also be affected by similar issues. Although this problem was discovered because of implausible sizes of some erroneous calls, there is no reason to assume that calls with smaller SV sizes could not be affected, too. We are not aware of any method to verify whether SV calls are affected by this issue, except for direct visual inspection of sequence alignments. We therefore assume that any large SV call-set generated by Sniffles2 is likely to contain a certain proportion of mischaracterised SV calls. Thus, SV calls may require visual or experimental validation.

In our systematic quality control, we removed only Sniffles2 calls considered as highly likely problematic because of their large sizes. Although the maximum expected size of SVs in healthy humans cannot be estimated with certainty, we decided on a lenient threshold of 30 Mbp, guided by a 21.3 Mbp duplication in a healthy human adult, previously reported in the literature^43^. Only 23 variants were longer than 30 Mbp and subsequently removed (see below).

### Reference panel generation

The imputation reference panel was constructed from a combination of the long-read derived SV calls as well as short-read derived SNV and short InDel calls generated from the NYGC’s resequencing efforts of the 1000 Genomes project^11^.

The SV calls were generated as described above, and were filtered to remove variants that did not have the term PASS in the FILTER column of the *VCF* file, were present in just 1 individual (private singletons or doubletons), were annotated as IMPRECISE, were <50 bp or >30 Mbp, or had genotypes missing in >20% of samples. See **Supplementary Table 22** and **Supplementary Figures 4 and 5** for statistics on raw and post-QC SV calls. In total, 107,445 SVs remained in the imputation panel after QC.

To generate the short variant calls, we downloaded the sequences for all 3202 samples from the International Genome Sample Resource (IGSR; https://www.internationalgenome.org/data-portal/data-collection/30x-grch38), aligned them to GRCh38, and jointly called them using the Sentieon DNAseq pipeline^44^ according to GATK best practices^45^.

We then filtered out InDels that satisfied “INFO/QD < 2.0 || INFO/FS > 200.0 || INFO/ReadPosRankSum < −20.0 || INFO/SOR > 10.0” and SNVs with “INFO/QD < 2.0 || INFO/FS > 60.0 || INFO/MQ < 40.0 || INFO/MQRankSum < −12.5 || INFO/ReadPosRankSum < −8.0”. Next, we subset the filtered, jointly called dataset to the 888 samples with long-read data and filtered out any variants present in <2 individuals.

The filtered long-read-derived SV callset was then merged using bcftools with the filtered short-read-derived short-variant callset, resulting in an unphased *VCF* file with sporadic missingness. Next, these sporadically missing genotypes were imputed using Beagle4^46^ and phased using Beagle5^47^, resulting in an imputation-ready reference panel *VCF* file. Default Beagle parameters were used except for the “--gtgl” flag instead of “--gl” during imputation because genotype likelihoods were absent for SVs. Beagle was run on chunks of 55,000 variants each, flanked by 3,000 variants up- and downstream.

### Reference panel characterisation

The generated reference panel of 888 unrelated 1000 Genomes Project individuals consisted of 45 million short variants (SNVs and InDels) and 107,445 SVs. In addition to stratification of the SVs by genomic region and variant types, we used frequency classes (MAF bins) for a more nuanced characterisation of SVs in the imputation panel (**Supplementary Table 4**).

We calculated the average counts of SV minor alleles per individual, stratified by superpopulation, SV type, and frequency class (**Supplementary Table 6**). Overall, individuals from the African superpopulation had the most diverse SVs, carrying on average 18,822 minor SV alleles, while individuals with East Asian ethnicities were least diverse, showing on average 14,792 SV minor alleles.

We analysed how SVs were shared across populations, stratified by SV type, frequency class, and region type. We counted SVs exclusive to specific populations and SVs shared between either two, three, four, or all five ancestry groups (**Supplementary Table 7**).

### Functional annotation of SVs

This section describes the annotation of SVs as shown in **Supplementary Table 5**.

For the purposes of filtering and functional interpretation of association results, SVs mapping to significant loci were annotated with the GIAB *genome stratification files*^13^ by intersecting bed files containing SV coordinates with genome stratification bed files (intersect -a ${SVbed} -b ${genome_stratification.bed} -c} and merging the genome stratification labels overlapping with SVs into a single column (column *GIAB_annotations*).

SnpEff^14^ v5.1d was used to annotate genes and functional consequences based on canonical transcripts (*command:* snpEff/exec/snpeff GRCh38.105 ${vcf} -canon) and SnpSift^48^ v5.1d for extracting fields of interest from the annotated *VCF* file:

gunzip -c ${vcf_annotated} | snpEff/scripts/vcfEffOnePerLine.pl | snpEff/exec/snpsift extractFields - CHROM POS ID “ANN[*].GENE” “ANN[*].GENEID” “ANN[*].FEATURE” “ANN[*].FEATUREID” “ANN[*].BIOTYPE” “ANN[*].IMPACT” “ANN[*].EFFECT” “ANN[*].ERRORS”)

For each SV, all fields in the output were merged into one row and added to the table (columns starting with *SNPeff*).

For annotating GWAS catalogue variants, we used the *All associations v1.0.2* file^49^ called *gwas_catalog_v1.0.2-associations_e109_r2023-06-03.tsv*and SnpSift^50^ (*command:* snpEff/exec/snpsift gwasCat -db ${GWAScatalog} ${vcf}). We extracted the fields of interest with bcftools using the query:

-f “%ID %GWASCAT_TRAIT %GWASCAT_P_VALUE %GWASCAT_OR_BETA %GWASCAT_REPORTED_GENE %GWASCAT_PUBMED_ID\n

These columns were added to the table with the prefix *GWASCAT*.

In addition, we used the Ensembl BioMart functionality to export regions from the *Ensembl Regulation 109* database, including the datasets “Human Regulatory Features”, “Human miRNA Target Regions”, and “Human Other Regulatory Regions” (corresponding to PHANTOM regulatory region predictions). The files exported from BioMart were converted to the *bed* format and intersected with SVs (intersect -a ${SVbed} -b ${genome_stratification.bed} -loj). For each SV, the annotations were added to the table as “regulatory_feature”, “predicted_regulatory_feature”, and “targeted_by_miRNA”, each with the prefix *Ensembl*.

### Generating a reduced panel for SV imputation

We created a backbone for imputation by using short variants available in the UK Biobank genotype data. To this end, the genomic positions of 805,426 directly genotyped UK Biobank variants were lifted over from the version GRCh37 to the GRCh38 human genome assembly using the DNAnexus pipeline^51^ based on Picard’s^40^ LiftoverVcf tool. 803,700 (99.8%) variants were successfully lifted over, 764,685 of which were SNVs. 702,464 of 764,685 (91.9%) of the genotyped SNVs could be matched to SNVs in our reference panel, serving as the basis for imputation into UKB (see below).

To facilitate imputation of SVs, we generated a reduced panel by removing millions of short variants not directly required for this purpose. The full reference panel containing ∼45M short variants and 107,445 SVs was reduced to include only the 702,464 SNVs directly genotyped in UKB, the 107,445 SVs, and 77,350 randomly selected short variants. The 77,350 random short variants (of which 57,733 were SNVs) were imputed along the SVs for benchmarking purposes. The filtering of the panel *VCF* file was performed using ‘awk’, retaining preannotated genotyped SNVs and SVs and a fraction of additional variants selected using awk’s *rand()* function. All the imputation steps described in the present study were performed using the reduced panel.

### Leave-one-out imputation performance

To assess whether the reduced reference panel is suitable for imputation, we performed leave-one-out cross-validation analyses: We excluded one individual from the panel and imputed SVs for this individual using the panel of the remaining 887 samples, applying exactly the same pipeline and settings as those later used for SV imputation into UK Biobank (see below). Then we compared the imputed SV genotypes to the sequenced genotypes of the excluded individual. To this end, we computed genotype concordance, non-reference concordance^52^, minor allele genotype concordance, and *r^2^_imp_* ^53^ from allele probabilities produced by Beagle v5.4. We repeated this leave-one-out analysis for each of the 888 individuals. Next, we computed individual-level mean concordances from SVs stratified by MAF, variant, and region class, and aggregated the metrics across superpopulations (**Supplementary Table 8**).

We also calculated the variant-level concordance and *r^2^_imp_* as aggregated imputation performance metrics for SVs (**Supplementary Table 9**). Across all 107,445 analysed SVs, the mean genotype concordance was 0.940 and *r^2^_imp_* =0.538. However, aggregate metrics are not meaningful for the interpretation of the overall imputation quality. Instead, the imputation quality needs to be stratified by SV type, MAF, and genomic region.

The interpretability of the concordance metric is limited in the presence of rare variants. For a rare allele *m*, most imputed genotypes have the genotype *MM* (*i.e.*, the homozygous genotype *MM* is most common). Consequently, a significant portion of genotypes may match between the ground truth and the imputed data purely by random chance. To better capture the imputation quality specifically of the minor alleles, the non-reference concordance metric was introduced in the literature^52^. Here, only the proportion of correctly imputed genotypes that contain non-reference alleles is assessed, ignoring homozygous reference alleles. However, a flaw in this metric becomes apparent when the reference allele corresponds to the minor allele. This issue can occur in genotype data, depending on the reference sequence and the composition of the cohort. In such cases, the intention of the non-reference concordance metric does not align with the actual computation since genotypes containing major alleles would be considered instead of genotypes containing minor alleles.

Consequently, this conceptual flaw skews the results of the non-reference concordance metric. For example, when calculating the metric on a sample-wise basis, the use of rare variants may lead to seemingly better performance compared to low-frequency variants (**Supplementary Table 8**). This misrepresents the fact that it is inherently more challenging to impute rare alleles accurately.

To mitigate this issue, we introduced the minor allele genotype concordance metric. This metric assesses the proportion of correctly imputed genotypes containing minor alleles, providing a more interpretable and intuitive measure compared to the non-reference concordance metric. By explicitly accounting for genotypes with minor alleles, this metric ensures a more accurate and interpretable evaluation of imputation performance, particularly for rare variants (**Supplementary Tables 8-9**).

### Preprocessing and imputation of SVs into UK Biobank

Imputation of SVs was performed in UK Biobank^19^ under the approved research proposal 57952.

#### Phasing of genotyped SNVs

After running liftover, we converted the output to the *VCF* format. Using PLINK2^54^ v2.00a3.8, we filtered the file to keep only SNVs (--snps-only) and aligned the files to a reference file (*Homo_sapiens.GRCh38.dna.primary_assembly.fa.gz* downloaded from Ensembl) using the “--ref-from-fa” parameter (to ensure the order of REF and ALT alleles remained compatible with the GRCh38 genome assembly). The resulting dataset of UKB-genotyped SNVs for 488,130 UKB individuals was then phased using SHAPEIT4 v4.2.2^55^. Each chromosome was phased separately using SHAPEIT4^56^ after splitting the chromosomes at the centromere region. We increased the number of Markov Chain Monte Carlo (MCMC) iterations for better phasing performance at cost of computational time, as recommended by the SHAPEIT4 authors (--mcmc-iterations 10b,1p,1b,1p,1b,1p,1b,1p,10m) and increased the number of conditioning neighbours in the Positional Burrows-Wheeler Transform (PBWT) to increase accuracy (--pbwt-depth 8). The resulting phased *VCF* files were concatenated per chromosome using the *concat* command of bcftools v1.16^57^.

#### Imputation of UK Biobank data

The imputation of SVs into UKB genotype data was performed with Beagle v5.4^47^. To this end, we converted our reduced panel from *VCF* format to Beagle’s *bref3* format. We could not impute the full set of UKB variants for each chromosome simultaneously due to limited computational resources (memory). Therefore, we divided the data for each chromosome in chunks of 10,000 individuals and then imputed each of these chunks independently. The imputation was conducted with Beagle v5.4 (file: *beagle.22Jul22.46e.jar*; downloaded from http://faculty.washington.edu/browning/beagle/beagle.html) using 32 cores (nthreads=32) and the genetic maps provided by the Beagle authors (map=”plink.chr${chr}.GRCh38.map”)^58^. We applied parameters to produce genotype and allele probabilities (gp=true, ap=true). Subsequently, the files per chromosome containing the imputed data for 10,000 individuals each were merged using the merge command of bcftools with the option *–merge none*. Finally, *r^2^_imp_* scores were re-computed based on all individuals, as outlined in the literature^53^ (**Supplementary Tables 10-11**).

### SV imputation verification using UK Biobank whole-genome sequencing

As a proof-of-principle verification, we used the recently published short-read whole-genome sequencing (WGS) data from UKB^18^ to detect the Sniffles2.DEL.3639MF deletion in UKB individuals. The read coverage was extracted from the population-level *VCF* files (UKB field 24310) by adding up the local allelic depth numbers (‘LAD’ field in the *VCF* files). We saw a clear drop in read coverage in the SV region chr16:75395953-75396795 in individuals that had one or both alleles deleted (**Supplementary Figure 7**). We compared the mean coverage at the SV site to the mean coverage on the flanking regions to detect the deletion genotype in each UKB individual. The individuals clustered very well into three groups, corresponding to 0, 1 or 2 deleted alleles (see **Supplementary Figure 8**). We compared this genotype to the imputed genotype for 485,129 samples present in both the WGS and imputed datasets. Excluding 1553 individuals for which the WGS calls were not confident (the ratio of coverage at and around Sniffles2.DEL.3639MF was in between the “no deletion” and “one allele deleted” clusters), the genotypes coincided for 98.7% of individuals (477086 out of 483576), exceeding our accuracy estimations based on the leave-one-out analysis (concordance of 92.3%). We only used this simple read coverage method to verify one imputed SV since it requires a significant manual effort and can only be used to detect deletions in non-repetitive areas of the genome. A full-scale SV calling from the UKB short-read WGS data is beyond the scope of this study.

### Genome-wide association study utilising imputed structural variants in UK Biobank

#### UK Biobank sample selection for association analyses

We performed SV genome-wide association analyses (SV-WAS) in UKB participants of European ancestry. For the selection of samples with “White European” ancestry, we adhered to a strategy previously established in the literature^59,60^. In summary, we applied k-means clustering (with k=6) on the first two genetic principal components from the principal component analysis (PCA) results provided by UKB (“ukb_sqc_v2_pca.txt”). We excluded individuals who had withdrawn consent or did not map to the European cluster (**Supplementary Figure 9**). We identified 171 duplicate pairs among the selected samples using KING v2.3.0 (--related). For each pair, we excluded the sample with the most missing phenotypes of interest. In cases where no significant differences were found, we randomly selected a sample from the pair to exclude. This resulted in a dataset of 455,589 samples for association analyses.

#### SV-wide association studies in UK Biobank

SV-WASs and SV-based pQTLs were calculated in UKB using imputed SV dosages and a linear mixed model, incorporating a leave-one-chromosome-out whole-genome regression model to account for population stratification as implemented in Regenie v3^61^. The effective number of individuals included in each analysis depended on the availability of phenotype and protein-level measurements. The number varied from 363K to 454K for quantitative and binary phenotypes (see Supplementary Tables 12 and 13) and was 48,205 for pQTL analyses. We computed the ancestry principal components separately for each phenotype set of individuals and for the pQTL set of individuals using PLINK2 with parameters ‘--pca approx 20 --chr 1-22 --snps-only’ based on a pruned set of 255,369 directly genotyped UKB variants. The pruning was also done with PLINK2 using ‘–maf 0.005 --indep-pairwise 200kb 0.5‘ on the set of all directly genotyped UKB variants.

This set of 255,369 pruned variants was used in the first step of the Regenie run. The *VCF* files of imputed variants generated by Beagle were first filtered for SVs and then transformed to the PLINK2 format (pgen) using the PLINK2 command ‘--make-pgen --vcf $filtered_vcf dosage=DS’, with the dosage information kept. Regenie uses dosages for association analysis whenever they are available. The first and the second step of Regenie were run on each phenotype and each of the 1463 protein levels with the following settings: ‘--bsize 1000 --loocv --gz --lowmem --lowmem-prefix’ for the first step, and ‘--bsize 200 --firth --firth-se --approx --pThresh 0.05 --minMAC 3’ at the second step. In addition, the ‘--bt’ flag was used for binary phenotypes and ‘--qt’ otherwise. For all 19 quantitative phenotypes, the measurement at the first examination was used to maximise sample size. Quantitative phenotype values and protein levels were transformed with rank-based inverse normal transformation in R (‘function(x) qnorm((rank(x, na.last = “keep”) - 0.5) / sum(!is.na(x)))’) before association analyses. We used the following covariates: sex and genotype array (UKB data field 22000) as binary covariates, age, age^2^, and the first 20 genetic principal components as quantitative covariates. For lung function-related traits (FEV_1_, FVC, FEV_1_/FVC), ‘ever smoked’ (data field 20160) was added as a binary covariate. Note that the Regenie output includes the statistical attributes of the association test for each variant, together with the allele frequency and INFO score for the set of participants in the given association analysis.

In the exploratory SV-WAS, we used the standard threshold for genome-wide significance of p < 5×10^−8^. For the pQTL analyses, we applied Bonferroni correction for multiple testing on top of that genome-wide threshold, correcting for the number of tested protein levels (n=1463): *p* < 5×10^−8^ / 1463 = 3.4×10^−11^. We extracted 4,528 significant (*p*<5×10^−8^) SV associations in all the SV-WASs performed, which were all included in Supplementary Table 14. Filtering down by Regenie metrics INFO>0.7 and MAF>0.01, 3,858 SV associations remained, corresponding to 1,898 unique SVs.

A similar table for pQTL SV associations (Supplementary Table 15) includes 23,810 associations at the *p*<5×10^−8^ significance level. Using a Bonferroni corrected *p*-value threshold *p*<3.4×10^−11^ and the INFO>0.7, MAF>0.01 criteria, 10,518 significant SV associations remained (for 3,723 unique SVs) for 1,101 proteins.

Signal selection and conditional analyses

For each phenotype and protein level, the significant SVs (GWAS *p*-value<5×10^−8^; pQTL *p*-value<5×10^−8^/1463) were split into sets separated by a genomic distance ≥100kb. For each set, we performed conditional analyses: First, SV coordinates were lifted from GRCh38 to GRCh37, then the SV imputed data was merged with the standard UKB short-variant imputed genotype data (data field 22828) at the locus with ±500kb flanking regions on each side. Next, we ran an iterative conditional analysis on the merged dataset to select independent association signals, conditioning on the set of top variants and adding the identified independent variants in a stepwise manner, stopping when the *p*-value of the top variant reached >5×10^−6^ (> 5×10^−6^/1463 for pQTLs) or after five iterations. We considered the SV as an independent and novel signal if it appeared as a top variant in any iteration of the conditional analysis.

### Utilising SV information to identify novel disease-relevant genes (SV2G) and improve post-GWAS locus-to-gene (L2G) prioritisation

#### From identified SV associations to causal genes (SV2G)

We conducted the following steps to identify the putatively causal gene(s) in the associated regions where an SV overlapping a protein-coding gene was the top variant (see ‘SV2G’ in **Figure 1b**): a) Examination of the literature, b) cis-e/pQTL-based phenome-wide Mendelian randomisation (MR) analyses (using the TwoSampleMR package^62^), c) finemapping and colocalisation (coloc v5)^63^, for all genes implicated by the SVs at each locus. GCTA-COJO^64^ and LD pruning (*r*^2^<0.01) were used to select cis-e/pQTLs (within ±1Mbp of the gene’s transcription start site) as instrumental variables (IVs) for each exposure (list of exposures in **Supplementary Table 23a**). Where we identified a strong MR association between gene expression and a trait (e.g., between *MEGF6* expression and a lung-disease-relevant trait), we manually checked the regional Manhattan plots to ensure that there was strong evidence for local colocalisation between the cis-e/pQTL signals used as the MR instruments and the outcome/trait GWAS (examples in **Supplementary Figures 10-14**). The outcome GWASs (n=7,429; **Supplementary Table 23b**) consisted mostly of a manually curated list from IEU OpenGWAS^65^ but also from internally conducted GWASs on 44 endophenotypes and outcomes related to respiratory and/or fibrotic diseases^66,67^. A Bayesian method was used to fine-map each associated locus to a set of variants that – assuming the causal variant was also included in the analysis – contains the underlying causal variant with 95% probability^68^. We set the parameter *W* (*i.e.*, the variance of the prior distribution of effect sizes) to 0.04 (≈0.21^2^) in the approximate Bayes factor formula – which equates to a 95% belief that the absolute relative risk is 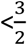 To estimate the probability that a single variant explains both the cis-e/pQTL signal and the signal in the trait/outcome GWAS, we manually inspected the regional Manhattan plots in addition to using the “coloc.abf” function^69^.

#### From SNV-based GWAS signals to causal genes utilising SV information (L2G)

To systematically analyse the contribution of SV information to post-GWAS locus-to-gene (see ‘L2G’ in **Figure 1b**) prioritisation approaches, we utilised the list of independent associations identified by the most recent GWAS of lung function carried out by Shrine *et al*^20^ and the table of implicated genes (consisting of genes implicated by various post-GWAS methods such as e/pQTL association, PoPS^21^, and rare respiratory disease causal genes) for each associated locus (Column AB in **Supplementary Table 18**). First, we lifted over the SNV coordinates reported by Shrine *et al.* (in GRCh37) to GRCh38 positions. We then looked for SVs that were (i) within 500 kb of the independent SNVs reported by Shrine *et al.*, (ii) significantly associated (*p*<5×10^−8^ in our UKB-based SV-WAS) with the primary lung function measure reported by Shrine *et al*^20^ (e.g. FEV_1_/FVC for rs2355210), and (iii) which spanned protein-coding genes (as defined by GENCODE v43). As a pragmatic approach to focus on the SVs likely to be real and functional, we restricted the analyses to a set of well-imputed SVs (INFO metric >0.7) not mapping to ‘difficult’ regions.

## Author contributions

Study conception and design: ZD, JS, SO, AME, BN, JNJ, NP, JK. Data governance/infrastructure, statistical analysis, and/or interpretation: BN, JS, SO, DD, AME, TFMA, CM, JCBL, BAB, CB, LS, SM, AK, JHL, GMB, IB. Preparation of the manuscript: ZD, AME, BN, TFMA, DD, SO, JS, JKP, JHL, GMB, JA, IB. All authors (incl. all under the ‘gCBDS’ banner) have critically reviewed and approved the final version of this paper, including the authorship statement.

## ‘Boehringer Ingelheim – Global Computational Biology and Digital Sciences’ authors

Johann de Jong, Fidel Ramirez, Frank Li, Thomas Kandl, James Cai, George Okafo.

**Address:** Global Computational Biology and Digital Sciences (gCBDS), Boehringer Ingelheim Pharma GmbH & Co. KG, Biberach an der Riss, Germany.

## Data availability

Raw SV calls, the long-read sequencing-based SV imputation panel, and the SV summary statistics from 32 SV-wide association studies are available through the OpnMe initiative of Boehringer Ingelheim GmbH (https://opnme.com/genomiclens). The raw long-read sequencing data (FASTQ files) for the 1000 Genomes Project samples included in this study are accessible via the European Nucleotide Archive under accession number PRJEB89727 (https://www.ebi.ac.uk/ena/browser/view/PRJEB89727). The dataset analysed here constitutes a subset of this broader collection. Imputed SVs for UK Biobank participants will be made available through the UKB Research Analysis Platform (RAP).

## Ethics declarations

This research has been conducted using the UK Biobank Resource under Application Number 57952.

## Acknowledgements

This research has been conducted using the UK Biobank, a major biomedical database (www.ukbiobank.ac.uk). We thank all UK Biobank and 1000 Genomes Project participants, without whom this project would not have been possible.

We express our gratitude to the MARVL Initiative – a collaboration between the Research Institute of Molecular Pathology (IMP), BI X and gCBDS (Boehringer Ingelheim). In particular: Siegfried Schloissnig, Klaus Ehrlinger, Julien Charest, Mila Asparuhova and Patrick Hüther for sequencing the samples.

We thank Gilean McVean and Jan Korbel for guidance during the project and providing critical feedback on the manuscript. We would like to thank the anonymous reviewers of the first version of this manuscript.

