## Supplementary Figures and Tables for "Imputation of structural variants using a multi-ancestry long-read sequencing panel enables identification of disease associations"

### Supplementary Figures 1-14

#### Supplementary Figure 1. Distribution of read statistics stratified by sequencing run

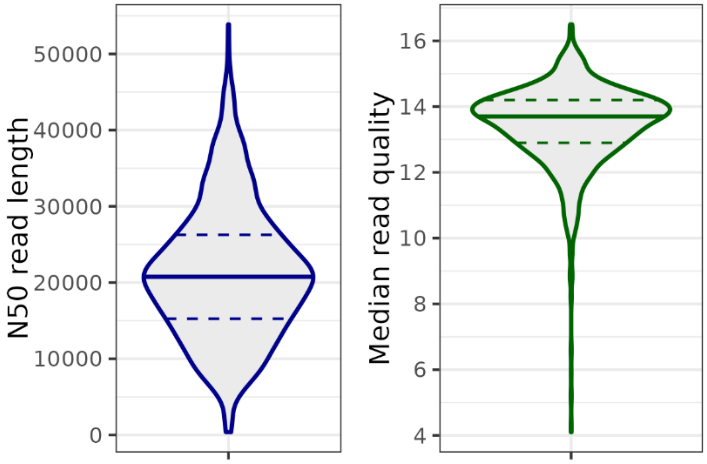


***Supplementary Figure 1: Distribution of read statistics by sequencing run.*** *Violin plots visualise the N50 read length per sequencing run and the median per run Phred read quality, as reported by NanoStat (see Supplementary Table 1). Some samples were sequenced in multiple runs due to insufficient coverage or quality in initial attempts, which contributes to the lower tails of the distributions.*

Supplementary Figure 2: Mean and median coverages in samples used for the imputation panel stratified by superpopulation.

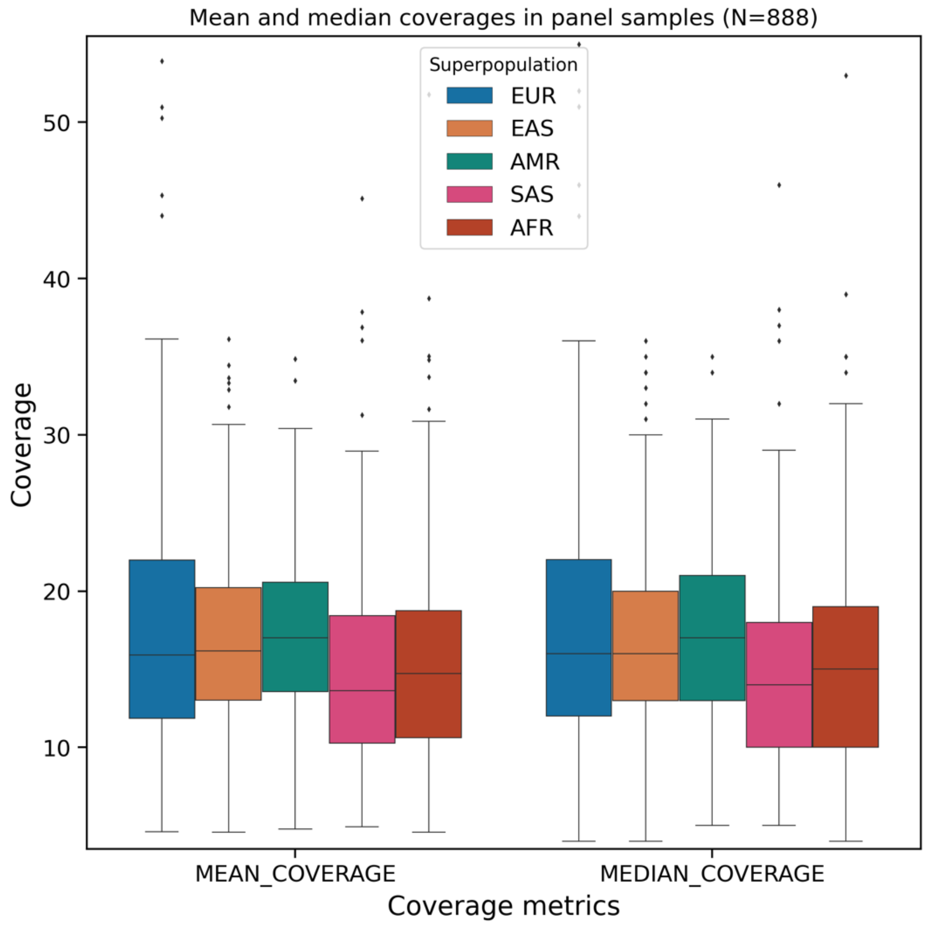


***Supplementary Figure 2: Mean and median coverages in samples used for the imputation panel stratified by superpopulation.*** *Variability was observed in the coverage metrics per sample, with mean coverage between 4.6x and 53.9x following sample quality control and application of default Picard filters. The median coverage per sample was within a 4x to 55x range. The distributions of sequencing coverages across different superpopulation groups were in the same range.*

#### Supplementary Figure 3. SV calling benchmarks vs short-read based SV callset.

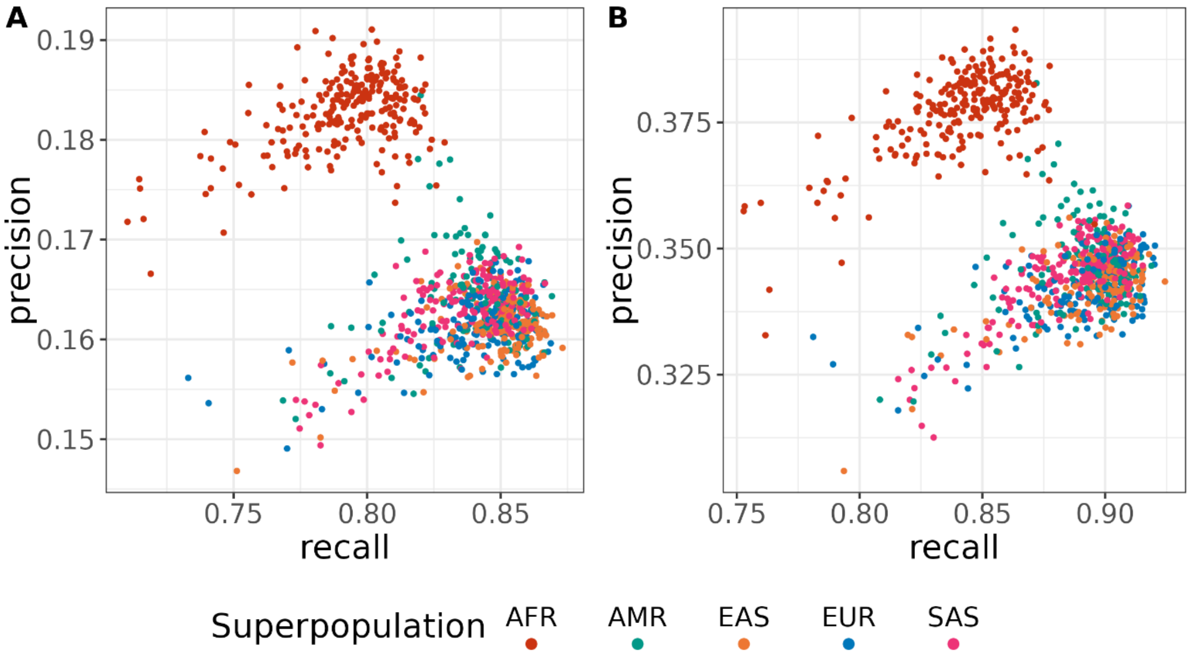


***Supplementary Figure 3: SV calling benchmarks vs short-read based SV callset.*** *Recall and precision rates for 888 samples for SVs called from our long read Oxford Nanopore sequencing data, compared to the SV calls from Illumina high-coverage short-read data (NYGC) for the same individuals. Each point represents one individual, coloured by superpopulation. (A) Whole genome comparison. (B) Comparison excluding SVs in tandem repeat regions longer than 200 bases.*

#### Supplementary Figure 4: Sample-wise genotype missingness rates

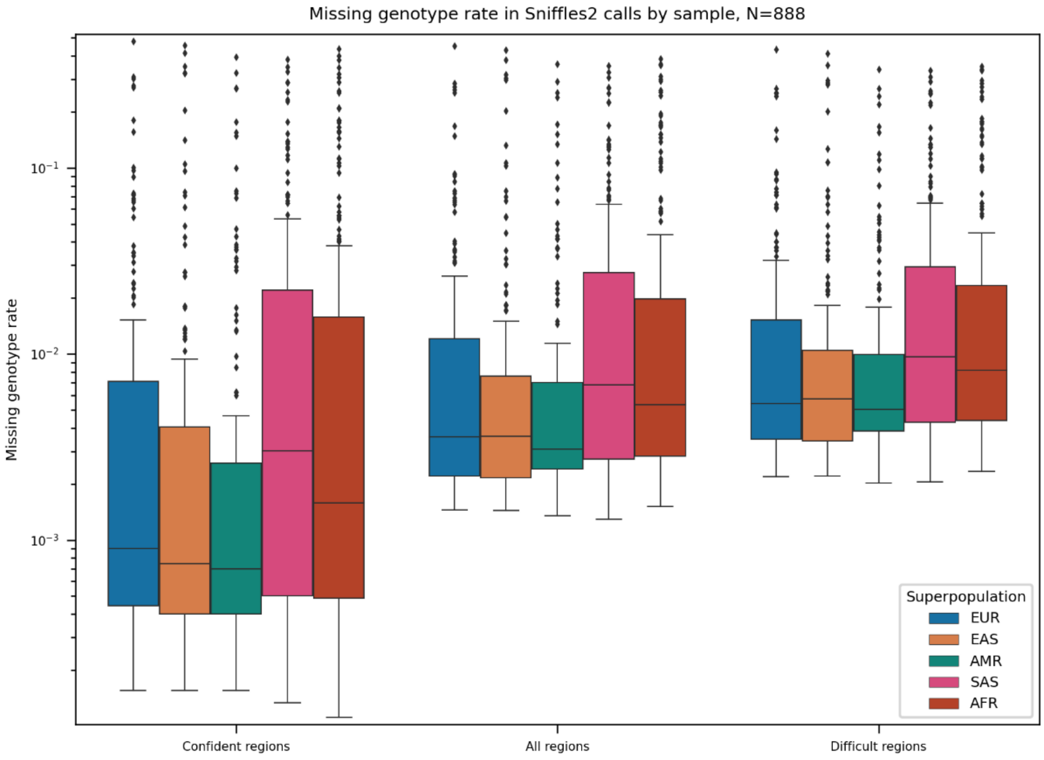


***Supplementary Figure 4: Sample-wise genotype missingness rates****. Missingness rates are stratified by superpopulation and region type (confident, all, difficult).*

#### Supplementary Figure 5: Genotype missingness rates stratified by SV type and region type

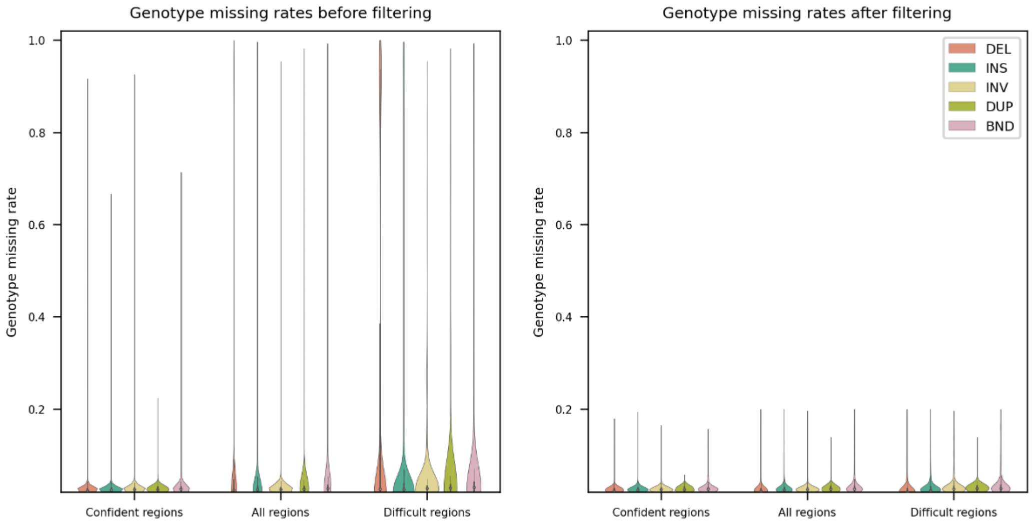


***Supplementary Figure 5: Genotype missingness rates stratified by SV type and region type.*** *Violin plots of missingness rate distributions before and after exclusion of SVs with high missingness rates ≥0.2 are shown. The missingness rates were computed in the 888 samples left after sample-wise QC.*

#### Supplementary Figure 6. Dependence of SV imputation quality and allele frequency on SV length

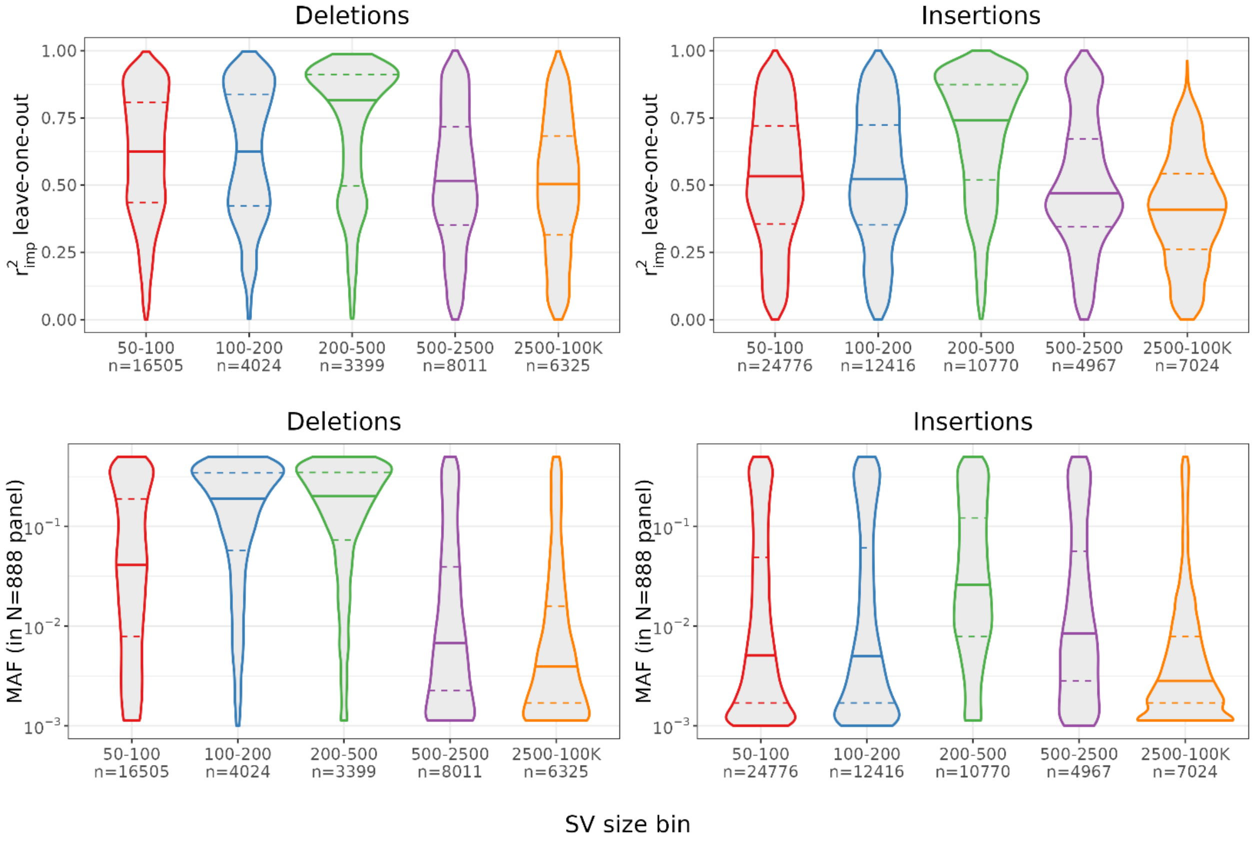


***Supplementary Figure 6: Dependence of SV imputation quality and allele frequency on SV length.*** *The leave-one-out imputation quality $r^2_{imp}$ and imputation panel MAF were summarised across five SV size bins. Deletions and insertions are included; inversions and duplications are not shown due to low counts that preclude stable binned estimates. The observed association between imputation quality and SV size is primarily driven by the length-dependent MAF in the imputation panel.*

#### Supplementary Figure 7: Drop in read coverage at the deletion site

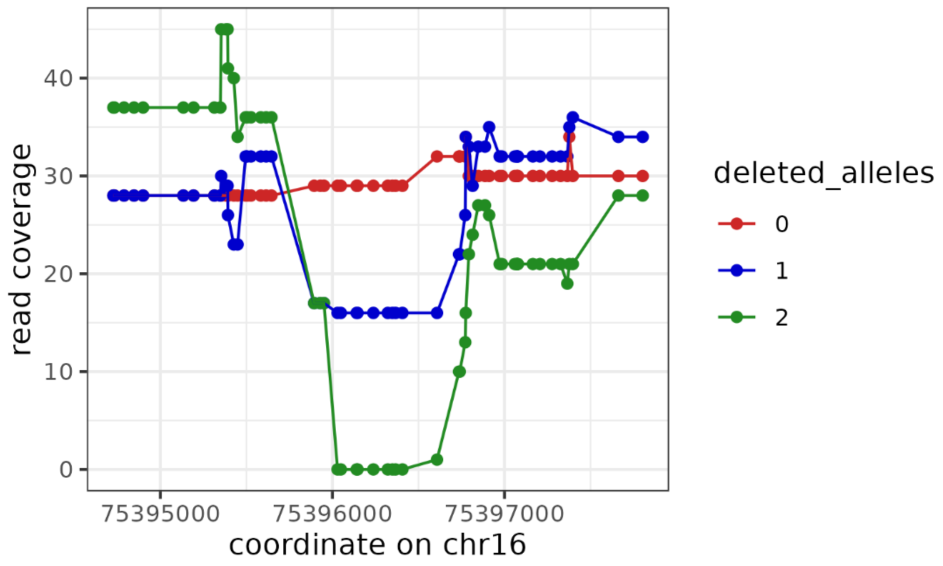


***Supplementary Figure 7: Drop in read coverage at the deletion site.*** *Examples of read coverage in the region containing the Sniffles2.DEL.3639MF deletion for 3 UKB samples – with 0,1 and 2 alleles deleted respectively. There is a clear drop in coverage in chr16:75395953-75396795.*

#### Supplementary Figure 8: Clustering of WGS read coverage values compared to the imputed SV genotypes

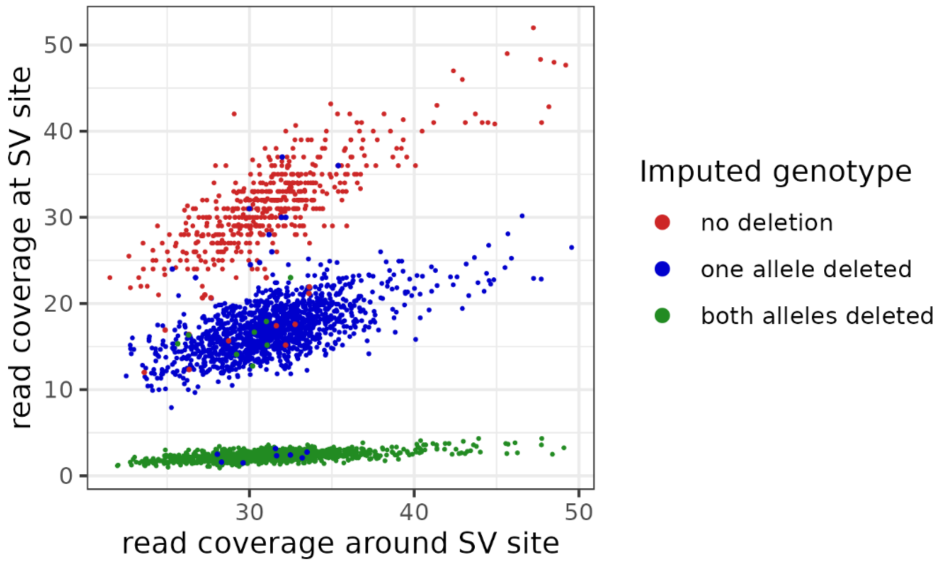


***Supplementary Figure 8. Clustering of WGS read coverage values compared to the imputed SV genotypes.*** *Scatter plot of WGS read coverage at the Sniffles2.DEL.3639MF deletion site vs that around the site for 3,000 randomly selected UKB individuals. Each point on the scatter plot corresponds to one individual, its colour is defined by the imputed genotype as shown in the legend. The 3 clusters fit very well the imputed genotypes with only 1.3% of mismatches across the entire UKB cohort.*

#### Supplementary Figure 9: Sample selection by k-means clustering

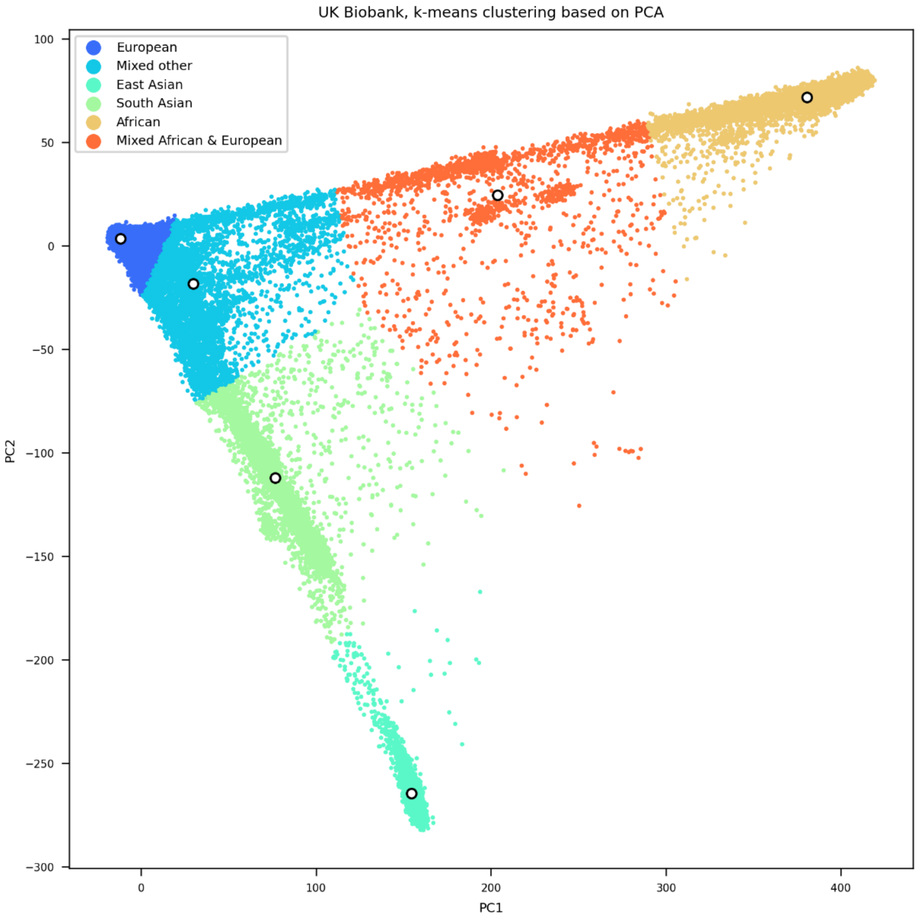


***Supplementary Figure 9: Sample selection by k-means clustering.*** *White circles represent cluster centres. Samples within the European ancestry cluster were selected for the genome-wide association studies.*

#### Supplementary Figure 10: Plots of the *SLC1A2* region in Smoking behaviour GWASs in UK Biobank

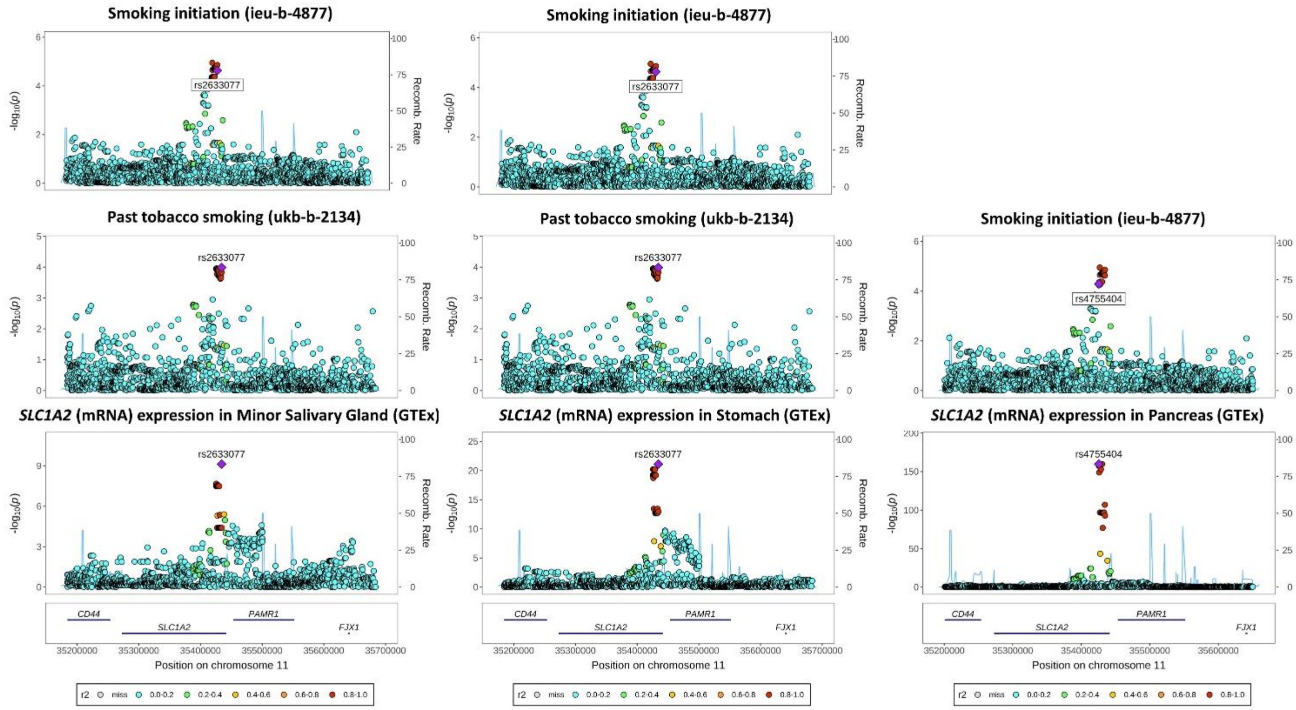


***Supplementary Figure 10: Evidence from other omics studies linking SLC1A2 to smoking behaviour-related traits**. Region plot showing strong local colocalisation between SLC1A2 (mRNA) expression in various GTEx tissues and smoking behaviour-related traits including smoking initiation (IEU Open GWAS ID: ieu-b-4877) – with the calculated colocalisation posterior probability (coloc H4) being ≥90% for all MR associations. Plot generated using LocusZoom. Full phenome-wide MR results for SLC1A2 (mRNA) expression are available in **Supplementary Table 25**. General information on the MR exposures and the outcome GWASs are available in **Supplementary Tables 23a-b**.*

#### Supplementary Figure 11: Plots of the *CFDP1* region in $FEV_1$/FVC<0.7 and PEF GWASs in UK Biobank

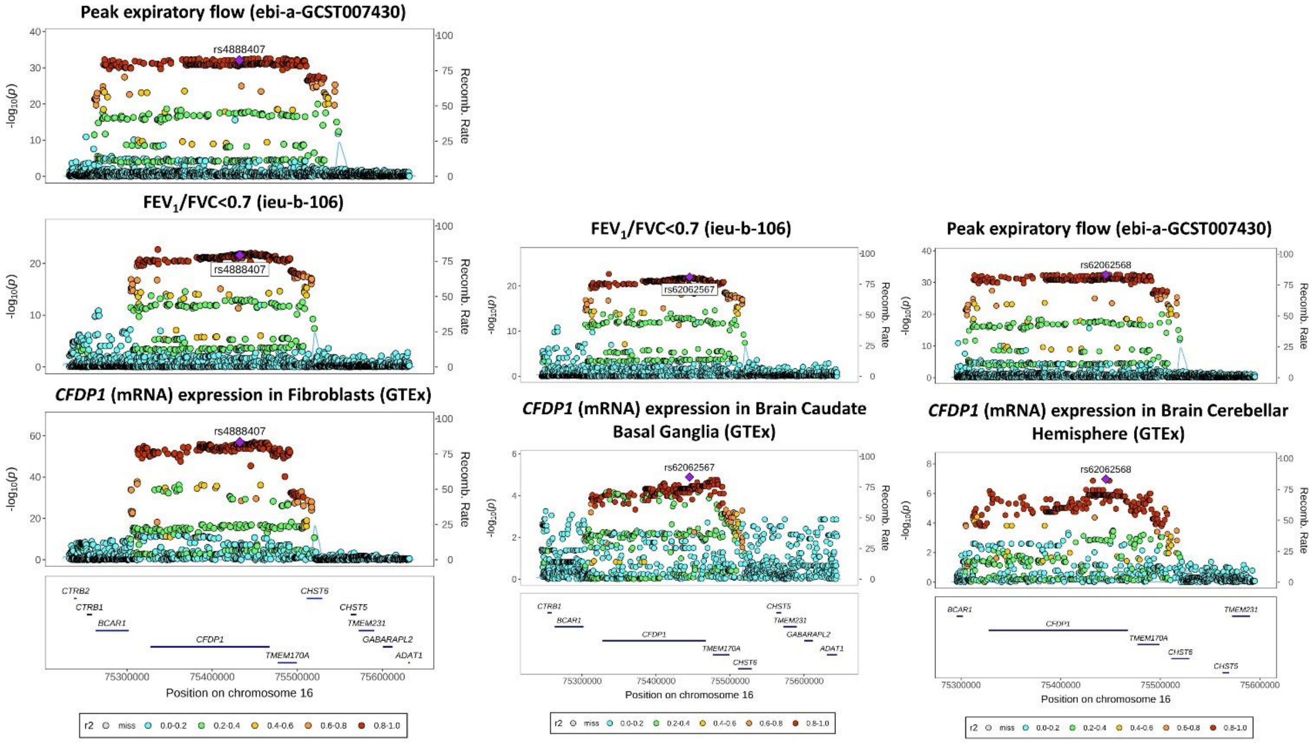


***Supplementary Figure 11: Plots of the CFDP1 region in FEV1/FVC<0.7 and peak expiratory flow (PEF) GWASs in UK Biobank.*** *Region plot showing strong local colocalisation between CFDP1 (mRNA) expression in fibroblasts and various brain tissues (GTEx), and peak expiratory flow (PEF, IEU Open GWAS ID: ebi-a-GCST007430) and $FEV_1$/FVC<0.7 (IEU Open GWAS ID: ieu-b-106). The calculated colocalisation posterior probability (coloc H4) was ~90% for all analyses. Plot generated using LocusZoom. Full phenome-wide MR results for CFDP1 (mRNA) expression are available in* ***Supplementary Table 19****. General information on the MR exposures and the outcome GWASs are available in* ***Supplementary Tables 23a-b****.*

#### Supplementary Figure 12: Plot of the *MEGF6* region in $FEV_1$/FVC GWAS in UK Biobank

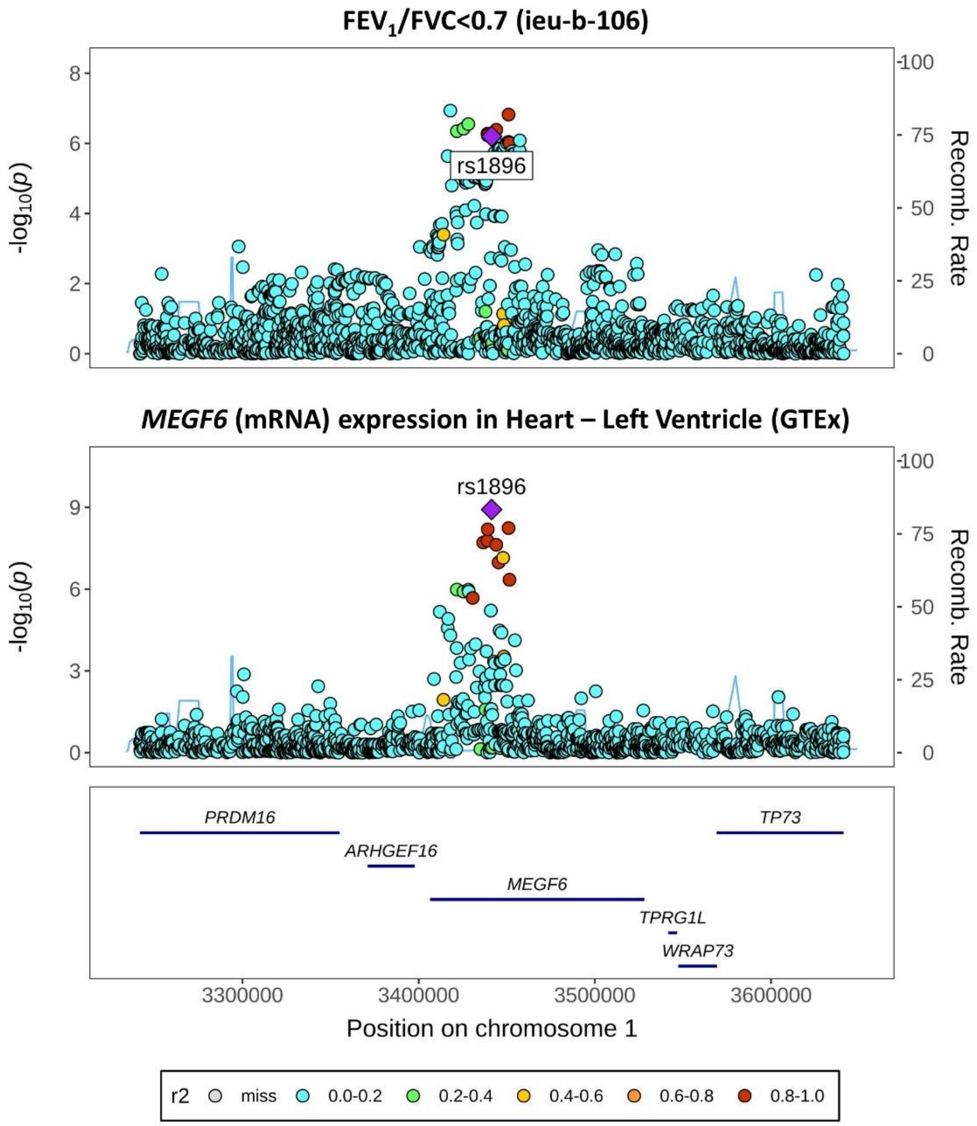


***Supplementary Figure 12: Plot of the MEGF6 region in the $FEV_1$/FVC GWAS in UK Biobank**. Region plot showing strong local colocalisation between MEGF6 (mRNA) expression in heart tissue (GTEx) and FEV1/FVC<0.7 (IEU Open GWAS ID: ieu-b-106) – a clinically used diagnostic criterion for chronic obstructive pulmonary disease. The calculated colocalisation posterior probability (coloc H4) was ~100%. There are also sQTLs for MEGF6 in this region (and not for the other genes nearby). Plot generated using LocusZoom. Full phenome-wide MR results for MEGF6 (mRNA) expression are available in **Supplementary Table 20**. General information on the MR exposures and the outcome GWASs are available in **Supplementary Tables 23a-b**.*

#### Supplementary Figure 13: Plots of the *AAGAB* region in lung function GWASs in UK Biobank

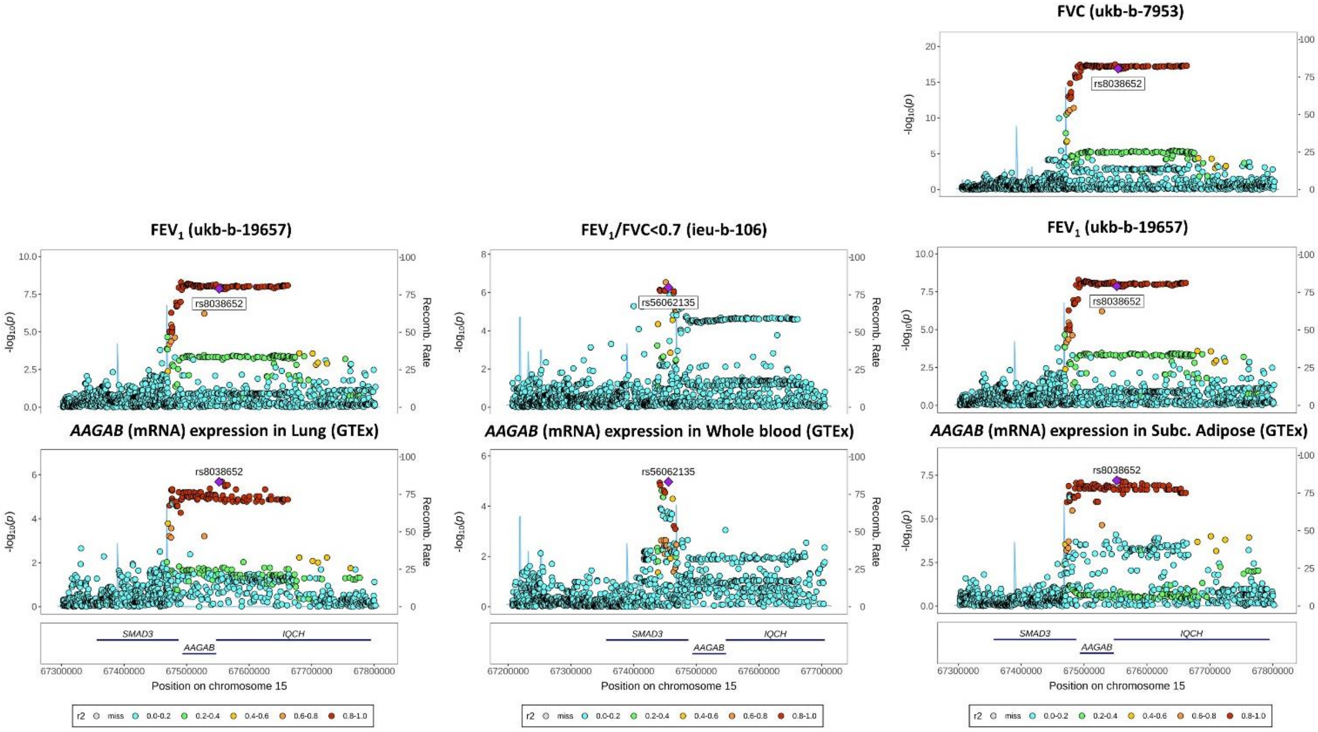


***Supplementary Figure 13: Plots of the AAGAB region in lung function GWASs in UK Biobank.*** *Region plots showing strong local colocalisation between AAGAB (mRNA) expression in various tissues including lung (GTEx) and various lung function measures including FEV$_1$/FVC<0.7 (IEU Open GWAS ID: ieu-b-106). The calculated colocalisation posterior probability (coloc H4) was >90% for all MR associations. Plot generated using LocusZoom. Full phenome-wide MR results for AAGAB (mRNA) expression are available in **Supplementary Table 21**. General information on the MR exposures and the outcome GWASs are available in **Supplementary Tables 23a-b**.*

#### Supplementary Figure 14: Evidence from eQTL-based Mendelian randomisation and colocalisation analyses linking *FLI1* expression to lung function and respiratory disease

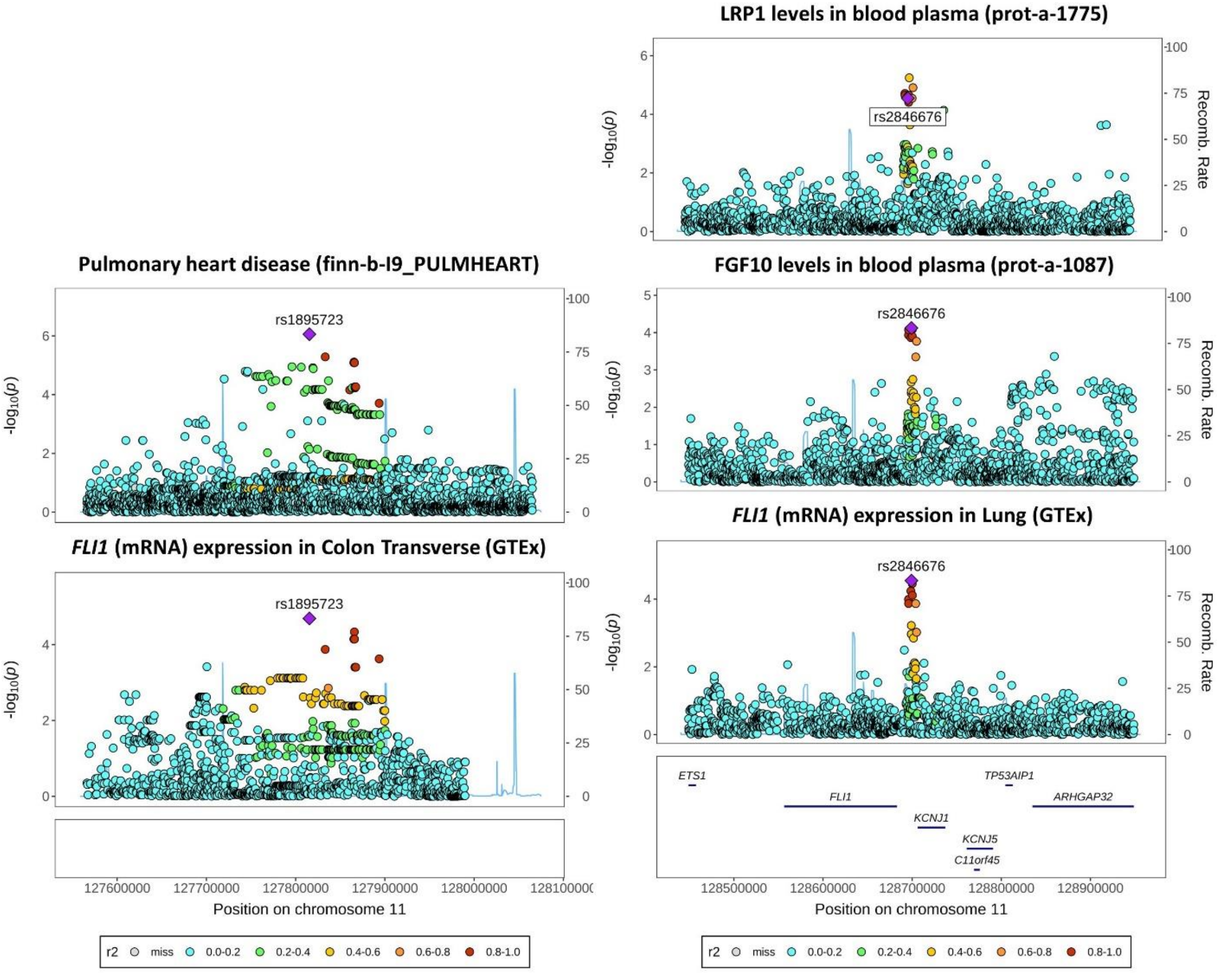


***Supplementary Figure 14: Evidence from other omics studies linking FLI1 to lung function and respiratory disease***. *Region plot showing strong local colocalisation between FLI1 (mRNA) expression in GTEx lung/smooth-muscle-containing tissues and (i) pulmonary heart disease risk (IEU Open GWAS ID: finn-b-I9_PULMHEART), (ii) LRP1 levels (IEU Open GWAS ID: prot-a-1775), and (iii) FGF10 levels (IEU Open GWAS ID: prot-a-1087) – with the calculated colocalisation posterior probability (coloc H4) being ≥80% for all three MR associations. Plot generated using LocusZoom. Full phenome-wide MR results for FLI1 (mRNA) expression are available in* ***Supplementary Table 24***. *General information on the MR exposures and the outcome GWASs are available in* ***Supplementary Tables 23a-b***.

### Supplementary Tables 3-4, 6-11 and 22

(Supplementary Tables 1-2, 5, 12–21, and 23–25 are provided in a separate Excel file.)

#### Legend for Supplementary Table 2

For the content of Supplementary Table 2 (Table of Picard WGS alignment performance statistics), please see the separate Excel file.

For a detailed description of Picard WGS alignment performance statistics, see the website https://broadinstitute.github.io/picard/picard-metric-definitions.html and the following table:

| FIELD | DESCRIPTION |
|---|---|
| **GENOME_TERRITORY** | The number of non-N bases in the genome reference over which coverage will be evaluated. |
| **MEAN_COVERAGE** | The mean coverage in bases of the genome territory, after all filters are applied. |
| **SD_COVERAGE** | The standard deviation of coverage of the genome after all filters are applied. |
| **MEDIAN_COVERAGE** | The median coverage in bases of the genome territory, after all filters are applied. |
| **MAD_COVERAGE** | The median absolute deviation of coverage of the genome after all filters are applied. |
| **PCT_EXC_MAPQ** | The fraction of aligned bases that were filtered out because they were in reads with low mapping quality (default is < 20). |
| **PCT_EXC_DUPE** | The fraction of aligned bases that were filtered out because they were in reads marked as duplicates. |
| **PCT_EXC_UNPAIRED** | The fraction of aligned bases that were filtered out because they were in reads without a mapped mate pair. |
| **PCT_EXC_BASEQ** | The fraction of aligned bases that were filtered out because they were of low base quality (default is < 20). |
| **PCT_EXC_OVERLAP** | The fraction of aligned bases that were filtered out because they were the second observation from an insert with overlapping reads. |
| **PCT_EXC_CAPPED** | The fraction of aligned bases that were filtered out because they would have raised coverage above the capped value (default cap = 250x). |
| **PCT_EXC_TOTAL** | The total fraction of aligned bases excluded due to all filters. |
| **PCT_1X** | The fraction of bases that attained at least 1X sequence coverage in post-filtering bases. |
| **PCT_5X** | The fraction of bases that attained at least 5X sequence coverage in post-filtering bases. |
| **PCT_10X** | The fraction of bases that attained at least 10X sequence coverage in post-filtering bases. |
| **PCT_15X** | The fraction of bases that attained at least 15X sequence coverage in post-filtering bases. |
| **PCT_20X** | The fraction of bases that attained at least 20X sequence coverage in post-filtering bases. |
| **PCT_25X** | The fraction of bases that attained at least 25X sequence coverage in post-filtering bases. |
| **PCT_30X** | The fraction of bases that attained at least 30X sequence coverage in post-filtering bases. |
| **PCT_40X** | The fraction of bases that attained at least 40X sequence coverage in post-filtering bases. |
| **PCT_50X** | The fraction of bases that attained at least 50X sequence coverage in post-filtering bases. |
| **PCT_60X** | The fraction of bases that attained at least 60X sequence coverage in post-filtering bases. |
| **PCT_70X** | The fraction of bases that attained at least 70X sequence coverage in post-filtering bases. |
| **PCT_80X** | The fraction of bases that attained at least 80X sequence coverage in post-filtering bases. |
| **PCT_90X** | The fraction of bases that attained at least 90X sequence coverage in post-filtering bases. |
| **PCT_100X** | The fraction of bases that attained at least 100X sequence coverage in post-filtering bases. |
| **HET_SNP_SENSITIVITY** | The theoretical HET SNP sensitivity. |
| **HET_SNP_Q** | The Phred Scaled Q Score of the theoretical HET SNP sensitivity. |

#### Supplementary Table 3: Sample counts in the imputation panel by superpopulation and population

| **Superpopulation** | Population code | Population description | Sample count |
|---|---|---|---|
| **European Ancestry (EUR)** | CEU | Utah residents (CEPH) with Northern and Western European ancestry | 26 |
| | FIN | Finnish in Finland | 34 |
| | GBR | British in England and Scotland | 36 |
| | IBS | Iberian populations in Spain | 38 |
| | TSI | Toscani in Italy | 30 |
| **East Asian Ancestry (EAS)** | CDX | Chinese Dai in Xishuangbanna, China | 36 |
| | CHB | Han Chinese in Beijing, China | 31 |
| | CHS | Han Chinese South | 40 |
| | JPT | Japanese in Tokyo, Japan | 26 |
| | KHV | Kinh in Ho Chi Minh City, Vietnam | 35 |
| **American Ancestry (AMR)** | CLM | Colombian in Medellin, Colombia | 42 |
| | MXL | Mexican Ancestry in Los Angeles, California | 27 |
| | PEL | Peruvian in Lima, Peru | 38 |
| | PUR | Puerto Rican in Puerto Rico | 37 |
| **South Asian Ancestry (SAS)** | BEB | Bengali in Bangladesh | 42 |
| | GIH | Gujarati Indians in Houston, TX | 28 |
| | ITU | Indian Telugu in the UK | 35 |
| | PJL | Punjabi in Lahore, Pakistan | 36 |
| | STU | Sri Lankan Tamil in the UK | 30 |
| **African Ancestry (AFR)** | ACB | African Caribbean in Barbados | 45 |
| | ASW | African Ancestry in Southwest US | 28 |
| | ESN | Esan in Nigeria | 43 |
| | GWD | Gambian in Western Division, The Gambia - Mandinka | 33 |
| | LWK | Luhya in Webuye, Kenya | 30 |
| | MSL | Mende in Sierra Leone | 34 |
| | YRI | Yoruba in Ibadan, Nigeria | 28 |

***Supplementary Table 3: Sample counts in the imputation panel by superpopulation and population.*** *Population codes and descriptions were obtained from the International Genome Sample Resource (IGSR) website* https://www.internationalgenome.org/data-portal/sample .

#### Supplementary Table 4: Variant counts in the imputation panel by variant type, frequency class, and region type

| Variant type | Frequency class | All regions | Confident regions | Difficult regions |
|---|---|---|---|---|
| **SNV** | Common | 12,351 | 9,349 | 3,002 |
| | Low frequency | 17,059 | 12,901 | 4,158 |
| | Rare | 27,467 | 21,328 | 6,139 |
| | All | 56,877 | 43,578 | 13,299 |
| **DEL** | Common | 16,145 | 4,665 | 11,480 |
| | Low frequency | 11,672 | 4,674 | 6,998 |
| | Rare | 10,642 | 5,884 | 4,758 |
| | All | 38,459 | 15,223 | 23,236 |
| **INS** | Common | 15,436 | 4,435 | 11,001 |
| | Low frequency | 18,054 | 6,605 | 11,449 |
| | Rare | 26,463 | 11,994 | 14,469 |
| | All | 59,953 | 23,034 | 36,919 |
| **INV** | Common | 58 | 44 | 14 |
| | Low frequency | 133 | 114 | 19 |
| | Rare | 5,538 | 4,887 | 651 |
| | All | 5,729 | 5,045 | 684 |
| **DUP** | Common | 12 | 6 | 6 |
| | Low frequency | 136 | 80 | 56 |
| | Rare | 460 | 332 | 128 |
| | All | 608 | 418 | 190 |
| **BND** | Common | 103 | 51 | 52 |
| | Low frequency | 818 | 354 | 464 |
| | Rare | 1,775 | 874 | 901 |
| | All | 2,696 | 1,279 | 1,417 |
| **All SVs** | | 107,445 | 44,999 | 62,446 |

***Supplementary Table 4: Variant counts in the imputation panel by variant type, MAF bin (frequency class), and region type.***

#### Supplementary Table 6: Sample-wise mean SV counts per superpopulation group

| | | Mean count per sample | | | | |
|---|---|---|---|---|---|---|
| **SV type** | **Frequency class** | **EUR** | **EAS** | **AMR** | **SAS** | **AFR** |
| **DEL** | Common | 7,415.52 | 7,358.81 | 7,483.90 | 7,490.40 | 8,553.57 |
| | Low frequency | 293.73 | 253.70 | 319.21 | 308.60 | 860.61 |
| | Rare | 32.98 | 37.88 | 31.85 | 45.78 | 74.56 |
| | All | 7,742.23 | 7,650.39 | 7,834.95 | 7,844.78 | 9,488.74 |
| **INS** | Common | 6,665.87 | 6,627.01 | 6,762.33 | 6,774.82 | 7,793.32 |
| | Low frequency | 379.04 | 337.54 | 421.49 | 396.32 | 1,231.98 |
| | Rare | 70.97 | 81.77 | 72.71 | 95.82 | 183.67 |
| | All | 7,115.88 | 7,046.32 | 7,256.53 | 7,266.97 | 9,208.97 |
| **INV** | Common | 23.84 | 24.35 | 25.39 | 24.33 | 23.64 |
| | Low frequency | 2.76 | 2.52 | 2.60 | 2.91 | 5.43 |
| | Rare | 9.38 | 10.11 | 8.53 | 23.08 | 24.61 |
| | All | 35.99 | 36.97 | 36.52 | 50.31 | 53.68 |
| **DUP** | Common | 3.61 | 3.38 | 3.57 | 3.81 | 3.40 |
| | Low frequency | 3.40 | 2.68 | 3.33 | 3.64 | 3.27 |
| | Rare | 1.57 | 1.22 | 1.49 | 2.50 | 2.17 |
| | All | 8.58 | 7.28 | 8.40 | 9.96 | 8.84 |
| **BND** | Common | 24.54 | 26.28 | 28.29 | 30.66 | 24.83 |
| | Low frequency | 18.66 | 19.74 | 20.31 | 26.56 | 26.39 |
| | Rare | 5.69 | 5.40 | 5.64 | 8.47 | 10.66 |
| | All | 48.88 | 51.42 | 54.24 | 65.69 | 61.87 |
| **All** | | 14,951.56 | 14,792.39 | 15,190.64 | 15,237.71 | 18,822.10 |

***Supplementary Table 6: Sample-wise mean SV counts per superpopulation group.*** *SV minor alleles and average counts of minor alleles were computed per superpopulation group and stratified by SV type and frequency class.*

#### Supplementary Table 7: SVs shared between superpopulation groups

| | | Ancestry-specific SV counts | | | | | Counts of SVs shared by ancestry groups | | | |
|---|---|---|---|---|---|---|---|---|---|---|
| **SV type** | **Frequency class** | **EUR** | **EAS** | **AMR** | **SAS** | **AFR** | **2 groups** | **3 groups** | **4 groups** | **5 groups** |
| **DEL** | Common | 0 | 0 | 0 | 0 | 2 | 90 | 180 | 643 | 15,231 |
| | Low frequency | 2 | 87 | 26 | 73 | 918 | 2,205 | 1,825 | 2,692 | 3,843 |
| | Rare | 324 | 860 | 240 | 919 | 2,556 | 3,406 | 1,644 | 608 | 85 |
| | All | 326 | 947 | 266 | 992 | 3,476 | 5,701 | 3,649 | 3,943 | 19,159 |
| **INS** | Common | 0 | 0 | 0 | 0 | 1 | 198 | 214 | 678 | 14,349 |
| | Low frequency | 3 | 147 | 27 | 150 | 1,968 | 4,544 | 3,012 | 3,347 | 4,852 |
| | Rare | 641 | 1,588 | 469 | 1,664 | 6,463 | 11,257 | 3,290 | 941 | 150 |
| | All | 644 | 1,735 | 496 | 1,814 | 8,432 | 15,999 | 6,516 | 4,966 | 19,351 |
| **INV** | Common | 0 | 0 | 0 | 0 | 0 | 0 | 0 | 4 | 54 |
| | Low frequency | 0 | 0 | 0 | 1 | 12 | 21 | 18 | 31 | 50 |
| | Rare | 34 | 68 | 32 | 283 | 731 | 3,457 | 805 | 119 | 9 |
| | All | 34 | 68 | 32 | 284 | 743 | 3,478 | 823 | 154 | 113 |
| **DUP** | Common | 0 | 0 | 0 | 0 | 0 | 0 | 0 | 0 | 12 |
| | Low frequency | 0 | 2 | 1 | 1 | 8 | 17 | 15 | 36 | 56 |
| | Rare | 25 | 46 | 22 | 56 | 97 | 122 | 59 | 24 | 9 |
| | All | 25 | 48 | 23 | 57 | 105 | 139 | 74 | 60 | 77 |
| **BND** | Common | 0 | 0 | 0 | 0 | 0 | 0 | 0 | 0 | 103 |
| | Low frequency | 0 | 1 | 1 | 0 | 29 | 57 | 91 | 202 | 437 |
| | Rare | 40 | 55 | 23 | 102 | 368 | 638 | 358 | 158 | 33 |
| | All | 40 | 56 | 24 | 102 | 397 | 695 | 449 | 360 | 573 |
| **All SVs** | | 1,069 | 2,854 | 841 | 3,249 | 13,153 | 26,012 | 11,511 | 9,483 | 39,273 |

***Supplementary Table 7: SVs shared between superpopulation groups.*** *Counts of SVs unique to samples in superpopulation groups, as well as counts of SVs shared by different numbers of superpopulations, stratified by SV type and frequency class.*

#### Supplementary Table 8: Sample-wise validation of imputation performance

| Variant type | Super-population | Frequency class | All regions: Concordance | All regions: Minor Allele Genotype Concordance | All regions: Non-reference Concordance | Confident regions: Concordance | Confident regions: Minor Allele Genotype Concordance | Confident regions: Non-reference Concordance | Difficult regions: Concordance | Difficult regions: Minor Allele Genotype Concordance | Difficult regions: Non-reference Concordance |
|---|---|---|---|---|---|---|---|---|---|---|---|
| SVs | EUR | Common | 0.854 | 0.718 | 0.760 | 0.940 | 0.885 | 0.903 | 0.819 | 0.657 | 0.708 |
| | | Low frequency | 0.981 | 0.407 | 0.798 | 0.990 | 0.633 | 0.861 | 0.976 | 0.3 | 0.774 |
| | | Rare | 0.997 | 0.141 | 0.625 | 0.997 | 0.202 | 0.597 | 0.996 | 0.089 | 0.646 |
| | EAS | Common | 0.847 | 0.696 | 0.747 | 0.933 | 0.867 | 0.891 | 0.811 | 0.633 | 0.695 |
| | | Low frequency | 0.981 | 0.292 | 0.782 | 0.989 | 0.498 | 0.839 | 0.975 | 0.209 | 0.761 |
| | | Rare | 0.996 | 0.085 | 0.580 | 0.997 | 0.126 | 0.534 | 0.996 | 0.049 | 0.613 |
| | AMR | Common | 0.850 | 0.712 | 0.756 | 0.936 | 0.878 | 0.897 | 0.815 | 0.651 | 0.704 |
| | | Low frequency | 0.980 | 0.407 | 0.790 | 0.988 | 0.599 | 0.839 | 0.975 | 0.311 | 0.771 |
| | | Rare | 0.997 | 0.134 | 0.628 | 0.997 | 0.195 | 0.6 | 0.996 | 0.081 | 0.65 |
| | SAS | Common | 0.836 | 0.686 | 0.734 | 0.924 | 0.856 | 0.88 | 0.800 | 0.625 | 0.681 |
| | | Low frequency | 0.978 | 0.326 | 0.762 | 0.987 | 0.522 | 0.82 | 0.972 | 0.24 | 0.74 |
| | | Rare | 0.995 | 0.104 | 0.534 | 0.996 | 0.136 | 0.474 | 0.995 | 0.072 | 0.581 |
| | AFR | Common | 0.794 | 0.653 | 0.690 | 0.888 | 0.806 | 0.83 | 0.756 | 0.6 | 0.641 |
| | | Low frequency | 0.951 | 0.482 | 0.679 | 0.958 | 0.59 | 0.706 | 0.946 | 0.408 | 0.665 |
| | | Rare | 0.993 | 0.205 | 0.477 | 0.993 | 0.254 | 0.445 | 0.992 | 0.151 | 0.506 |
| SNVs | EUR | Common | 0.961 | 0.919 | 0.933 | 0.979 | 0.958 | 0.965 | 0.906 | 0.807 | 0.836 |
| | | Low frequency | 0.994 | 0.741 | 0.878 | 0.995 | 0.798 | 0.911 | 0.990 | 0.591 | 0.763 |
| | | Rare | 0.998 | 0.319 | 0.579 | 0.998 | 0.337 | 0.611 | 0.997 | 0.27 | 0.468 |
| | EAS | Common | 0.951 | 0.897 | 0.917 | 0.968 | 0.934 | 0.947 | 0.898 | 0.79 | 0.825 |
| | | Low frequency | 0.992 | 0.580 | 0.823 | 0.993 | 0.634 | 0.857 | 0.988 | 0.437 | 0.701 |
| | | Rare | 0.997 | 0.166 | 0.454 | 0.997 | 0.18 | 0.485 | 0.996 | 0.123 | 0.34 |
| | AMR | Common | 0.957 | 0.912 | 0.927 | 0.975 | 0.95 | 0.958 | 0.902 | 0.802 | 0.833 |
| | | Low frequency | 0.991 | 0.689 | 0.839 | 0.993 | 0.745 | 0.875 | 0.986 | 0.538 | 0.715 |
| | | Rare | 0.998 | 0.303 | 0.588 | 0.998 | 0.323 | 0.622 | 0.997 | 0.244 | 0.463 |
| | SAS | Common | 0.951 | 0.900 | 0.917 | 0.969 | 0.939 | 0.949 | 0.896 | 0.789 | 0.821 |
| | | Low frequency | 0.992 | 0.660 | 0.843 | 0.993 | 0.714 | 0.877 | 0.988 | 0.522 | 0.725 |
| | | Rare | 0.996 | 0.239 | 0.477 | 0.997 | 0.256 | 0.508 | 0.996 | 0.185 | 0.364 |
| | AFR | Common | 0.915 | 0.848 | 0.868 | 0.931 | 0.88 | 0.896 | 0.864 | 0.752 | 0.782 |
| | | Low frequency | 0.964 | 0.665 | 0.724 | 0.966 | 0.69 | 0.748 | 0.960 | 0.579 | 0.64 |
| | | Rare | 0.994 | 0.371 | 0.478 | 0.994 | 0.389 | 0.501 | 0.993 | 0.306 | 0.39 |

***Supplementary Table 8: Sample-wise validation of imputation performance.*** *Mean sample-wise concordance per superpopulation, frequency class, and region type is shown. Concordances were computed using two variant classes: imputed SNVs and imputed SVs (combined across all SV types).*

### Supplementary Table 9: Validation of imputation performance by variant type

| | | All regions | | | | Confident regions | | | | Difficult regions | | | |
|---|---|---|---|---|---|---|---|---|---|---|---|---|---|
| **Variant type** | **Frequency class** | **Concordance** | Minor allele genotype concordance | Non-reference concordance | $r^2_{imp}$ | **Concordance** | Minor allele genotype concordance | Non-reference concordance | $r^2_{imp}$ | **Concordance** | Minor allele genotype concordance | Non-reference concordance | $r^2_{imp}$ |
| **SNV** | Common | 0.944 | 0.877 | 0.885 | 0.881 | 0.961 | 0.909 | 0.915 | 0.909 | 0.891 | 0.778 | 0.790 | 0.793 |
| | Low frequency | 0.985 | 0.609 | 0.614 | 0.696 | 0.986 | 0.644 | 0.649 | 0.717 | 0.980 | 0.500 | 0.504 | 0.632 |
| | Rare | 0.996 | 0.266 | 0.267 | 0.453 | 0.996 | 0.281 | 0.282 | 0.460 | 0.996 | 0.213 | 0.213 | 0.429 |
| | All | 0.981 | 0.501 | 0.505 | 0.619 | 0.986 | 0.523 | 0.527 | 0.632 | 0.967 | 0.430 | 0.434 | 0.575 |
| **DEL** | Common | 0.833 | 0.673 | 0.693 | 0.721 | 0.924 | 0.832 | 0.846 | 0.852 | 0.796 | 0.608 | 0.631 | 0.669 |
| | Low frequency | 0.969 | 0.351 | 0.367 | 0.554 | 0.979 | 0.499 | 0.515 | 0.644 | 0.963 | 0.252 | 0.268 | 0.493 |
| | Rare | 0.995 | 0.112 | 0.114 | 0.390 | 0.996 | 0.167 | 0.168 | 0.416 | 0.994 | 0.043 | 0.047 | 0.357 |
| | All | 0.919 | 0.420 | 0.434 | 0.579 | 0.968 | 0.473 | 0.482 | 0.620 | 0.887 | 0.385 | 0.402 | 0.552 |
| **INS** | Common | 0.832 | 0.655 | 0.696 | 0.718 | 0.921 | 0.830 | 0.846 | 0.858 | 0.797 | 0.584 | 0.636 | 0.661 |
| | Low frequency | 0.974 | 0.389 | 0.440 | 0.585 | 0.982 | 0.539 | 0.566 | 0.674 | 0.969 | 0.302 | 0.368 | 0.533 |
| | Rare | 0.995 | 0.128 | 0.132 | 0.400 | 0.996 | 0.174 | 0.177 | 0.422 | 0.995 | 0.089 | 0.094 | 0.381 |
| | All | 0.947 | 0.342 | 0.370 | 0.537 | 0.978 | 0.405 | 0.417 | 0.579 | 0.928 | 0.303 | 0.340 | 0.512 |
| **INV** | Common | 0.76 | 0.494 | 0.496 | 0.587 | 0.774 | 0.542 | 0.544 | 0.621 | 0.716 | 0.344 | 0.344 | 0.480 |
| | Low frequency | 0.972 | 0.206 | 0.206 | 0.494 | 0.972 | 0.225 | 0.225 | 0.511 | 0.974 | 0.095 | 0.095 | 0.386 |
| | Rare | 0.996 | 0.006 | 0.006 | 0.347 | 0.996 | 0.007 | 0.007 | 0.347 | 0.996 | 0.004 | 0.004 | 0.346 |
| | All | 0.993 | 0.016 | 0.016 | 0.353 | 0.994 | 0.016 | 0.016 | 0.353 | 0.990 | 0.013 | 0.013 | 0.350 |
| **DUP** | Common | 0.708 | 0.217 | 0.217 | 0.408 | 0.762 | 0.228 | 0.228 | 0.437 | 0.654 | 0.206 | 0.206 | 0.379 |
| | Low frequency | 0.972 | 0.159 | 0.159 | 0.455 | 0.974 | 0.251 | 0.251 | 0.502 | 0.969 | 0.027 | 0.027 | 0.389 |
| | Rare | 0.995 | 0.116 | 0.116 | 0.401 | 0.995 | 0.158 | 0.158 | 0.435 | 0.994 | 0.007 | 0.007 | 0.314 |
| | All | 0.984 | 0.127 | 0.127 | 0.413 | 0.988 | 0.177 | 0.177 | 0.448 | 0.976 | 0.019 | 0.019 | 0.338 |
| **BND** | Common | 0.744 | 0.233 | 0.234 | 0.478 | 0.760 | 0.251 | 0.251 | 0.504 | 0.729 | 0.215 | 0.217 | 0.454 |
| | Low frequency | 0.964 | 0.058 | 0.058 | 0.413 | 0.965 | 0.086 | 0.086 | 0.448 | 0.963 | 0.036 | 0.036 | 0.386 |
| | Rare | 0.994 | 0.025 | 0.025 | 0.368 | 0.994 | 0.036 | 0.036 | 0.379 | 0.994 | 0.014 | 0.014 | 0.357 |
| | All | 0.976 | 0.043 | 0.043 | 0.386 | 0.977 | 0.059 | 0.059 | 0.403 | 0.974 | 0.029 | 0.029 | 0.370 |
| **All SVs** | | 0.940 | 0.344 | 0.364 | 0.538 | 0.976 | 0.372 | 0.382 | 0.561 | 0.915 | 0.323 | 0.352 | 0.521 |

***Supplementary Table 9: Validation of imputation performance by variant type.*** *Mean imputation performance metrics (concordance and $r^2_{imp}$) from leave-one-out cross-validation analysis are shown, stratified by variant type, frequency class, and region type. In addition to SVs, validation metrics are shown for 56,877 imputed SNVs, which were randomly selected from the 1000 Genomes Project and included in the panel for validation purposes.*

#### Supplementary Table 10: Counts of imputed variants in UK Biobank data

| Variant type | Frequency class | All regions | Confident regions | Difficult regions |
|---|---|---|---|---|
| SNV | Common | 10,626 | 8,100 | 2,526 |
| | Low frequency | 6,793 | 5,001 | 1,792 |
| | Rare | 39,878 | 30,862 | 9,016 |
| | All | 57,297 | 43,963 | 13,334 |
| DEL | Common | 13,560 | 4,033 | 9,527 |
| | Low frequency | 6,748 | 2,104 | 4,643 |
| | Rare | 17,769 | 9,010 | 8,746 |
| | All | 38,077 | 15,147 | 22,916 |
| INS | Common | 12,839 | 3,864 | 8,975 |
| | Low frequency | 8,978 | 2,599 | 6,379 |
| | Rare | 37,468 | 16,421 | 21,026 |
| | All | 59,285 | 22,884 | 36,380 |
| INV | Common | 51 | 43 | 8 |
| | Low frequency | 166 | 138 | 27 |
| | Rare | 5,415 | 4,795 | 618 |
| | All | 5,632 | 4,976 | 653 |
| DUP | Common | 11 | 7 | 4 |
| | Low frequency | 91 | 64 | 27 |
| | Rare | 503 | 344 | 158 |
| | All | 605 | 415 | 189 |
| BND | Common | 78 | 38 | 40 |
| | Low frequency | 426 | 188 | 238 |
| | Rare | 2,154 | 1,040 | 1,111 |
| | All | 2,658 | 1,266 | 1,389 |
| All SVs | | 106,257 | 44,688 | 61,527 |

***Supplementary Table 10: Counts of imputed variants in UK Biobank data.*** *Only non-monomorphic variants (MAF>0) were used for computing counts and imputation metrics ($r^2_{imp}$). Frequency classes were determined using the MAF in the imputed data. All variant types except SNV refer to SVs.*

### Supplementary Table 11: Imputation quality in UK Biobank imputed variants

| Variant type | Frequency class | All regions | Confident regions | Difficult regions |
|---|---|---|---|---|
| **SNV** | Common | 0.937 | 0.961 | 0.862 |
| | Low frequency | 0.807 | 0.834 | 0.735 |
| | Rare | 0.472 | 0.481 | 0.439 |
| | All | 0.598 | 0.61 | 0.559 |
| **DEL** | Common | 0.803 | 0.915 | 0.756 |
| | Low frequency | 0.639 | 0.764 | 0.582 |
| | Rare | 0.391 | 0.440 | 0.340 |
| | All | 0.582 | 0.612 | 0.562 |
| **INS** | Common | 0.801 | 0.915 | 0.752 |
| | Low frequency | 0.660 | 0.782 | 0.610 |
| | Rare | 0.405 | 0.442 | 0.376 |
| | All | 0.529 | 0.561 | 0.51 |
| **INV** | Common | 0.695 | 0.715 | 0.587 |
| | Low frequency | 0.682 | 0.694 | 0.624 |
| | Rare | 0.307 | 0.307 | 0.308 |
| | All | 0.322 | 0.321 | 0.324 |
| **DUP** | Common | 0.567 | 0.602 | 0.504 |
| | Low frequency | 0.627 | 0.657 | 0.555 |
| | Rare | 0.340 | 0.364 | 0.287 |
| | All | 0.387 | 0.413 | 0.33 |
| **BND** | Common | 0.589 | 0.623 | 0.557 |
| | Low frequency | 0.555 | 0.566 | 0.546 |
| | Rare | 0.335 | 0.358 | 0.314 |
| | All | 0.378 | 0.397 | 0.361 |
| **All SVs** | | 0.533 | 0.545 | 0.523 |

***Supplementary Table 11: Imputation quality in UK Biobank imputed variants**. Imputation quality is represented by mean $r^2_{imp}$, stratified by variant type and frequency class, and is shown for all regions, confident regions, and difficult regions. Frequency classes were determined using the MAF in the imputed data. Note that all variant types except SNV refer to SVs.*

#### Supplementary Table 22: Genotype missingness rates by SV type and region type

| | SV type | All regions | Confident regions | Difficult regions |
|---|---|---|---|---|
| **Raw calls** | **DEL** | 0.166 | 0.023 | 0.233 |
| | **INS** | 0.086 | 0.025 | 0.119 |
| | **INV** | 0.031 | 0.027 | 0.057 |
| | **DUP** | 0.043 | 0.027 | 0.074 |
| | **BND** | 0.067 | 0.029 | 0.098 |
| **Post-QC calls** | **DEL** | 0.025 | 0.023 | 0.026 |
| | **INS** | 0.027 | 0.024 | 0.028 |
| | **INV** | 0.027 | 0.026 | 0.032 |
| | **DUP** | 0.028 | 0.027 | 0.029 |
| | **BND** | 0.031 | 0.028 | 0.033 |

***Supplementary Table 22: Genotype missingness rates by SV type and region type.*** *Mean genotype missingness rates in raw calls and after QC (removal of SVs with genotype missingness rate>0.2) are shown.*
